# Seeing Opportunity in Virtual Reality: A Rapid Review of the Use of VR as a Tool in Vision Care

**DOI:** 10.1101/2025.05.20.25327918

**Authors:** Kiana Masoudi, Madeline Wong, Danielle Tchao, Ani Orchanian-Cheff, Michael Reber, Lora Appel

**Affiliations:** OpenLab, Universtiy Health Network, Toronto, ON, CA; Library and Information Services, University Health Network, Toronto, ON, CA; Donald K Johnson Eye Institute, Krembil Fresearch Institute, University Health Network, Toronto, ON, CA; Department of Ophthalmology and Vision Sciences, University of Toronto, Toronto, ON, CA; INSERM UMR-S1329 STEP, Strasbourg, FR; School of Health Policy and Management, Faculty of Health, York University, Toronto, ON, CA; KITE, University Health Network, Toronto, ON, CA; General Internal Medicine, Michael Garron Hospital, Toronto, ON, CA

**Keywords:** Virtual Reality, Vision Disorders, Eye Care, Vision Therapy, Vision Screening, Visual Perception, Ophthalmology

## Abstract

**Background:** Virtual reality (VR) technologies have shown significant potential for diagnosing and treating vision-related impairments: This rapid review evaluates and characterizes the existing literature on VR technologies for diagnosing and treating vision-based diseases.

**Methods:** A systematic search was conducted across Ovid MEDLINE, Ovid Embase, the Cochrane Database of Systematic Reviews (Ovid), and the Cochrane Central Register of Controlled Trials (Ovid). Abstracts were screened using Rayyan QCRI, followed by full-text screening and data extraction. Eligible studies were published in peer-reviewed journals, written in English, focused on human participants, used immersive and portable VR devices as the primary intervention, and reported on the clinical effectiveness of VR for therapeutic, diagnostic, or screening purposes for vision or auditory-visual impairments. Various study characteristics, including design and participant details, were extracted, and the MMAT assessment tool was used to evaluate study quality.

**Results:** Seventy-six studies met the inclusion criteria. Among these, sixty-four (84.2%) were non-randomized studies exploring VR’s effectiveness, while twenty-two (15.8%) were randomized-controlled trials. Of the included studies, 38.2% focused on diagnosing, 21.0% on screening, and 38.2% on treating vision impairments. Glaucoma and amblyopia were the most commonly studied visual impairments.

**Conclusion:** The use of standalone, remotely controlled VR headsets for screening and diagnosing visual diseases represents a promising advancement in ophthalmology. With ongoing technological developments, VR has the potential to revolutionize eye care by improving accessibility, efficiency, and personalization. Continued research and innovation in VR applications for vision care are expected to further enhance patient outcomes.

**Systematic review registration:** PROSPERO CRD42023456214.

## Background

Virtual Reality (VR) technologies have emerged as increasingly valuable tools in healthcare, offering innovative applications in fields such as ophthalmology and multisensory processing. The ability of VR to engage both visual and auditory senses, which are crucial for spatial awareness and cognitive integration, provides a unique advantage in these contexts.

Virtual reality (VR) refers to an immersive technology that simulates a computer-generated environment, allowing users to perceive themselves as an active participant within this synthetic realm (Kouijzer et al. 2023). This immersive experience can be facilitated through a variety of display systems, including two-dimensional screens, stereoscopic 3D glasses, or more advanced head-mounted displays (HMDs), each offering varying levels of interaction and immersion (McMahan et al., 2006; McMahan et al., 2012).

HMDs are sophisticated VR devices that consist of a helmet or headgear fitted with screens that project images to each eye. Integrated position-tracking systems within these devices monitor the user’s gaze, enabling the real-time updating of visual content based on the viewer’s head movements. This results in a seamless and interactive virtual experience, which is often described as highly immersive (Bailenson, 2018). Over the past decade, the development of more affordable and accessible HMDs has significantly expanded their use (Fan et al., 2022; Jensen & Konradsen, 2018), transforming them into essential tools in both research and clinical applications (Kourtesis et al., 2020; Pottle, 2019). As a consequence, HMD-based VR has gained traction as a pivotal technology in the healthcare sector, including ophthalmology, where it supports therapeutic and diagnostic innovations (Freeman et al., 2020).

Beyond the overarching advantages of telehealth that VR offers, including increased accessibility to healthcare, improved efficiency for both healthcare providers and patients, and reduced costs—there are also substantial benefits specific to the immersive nature of this technology. VR’s unique capacity to fully immerse users in an interactive virtual environment lends it ecological validity for a wide range of clinical applications. Immersive VR, when combined with advanced technologies such as inertial measurement units (IMUs) and eye-tracking cameras, provides valuable insights into physiological measures such as eye movements (e.g., saccades, pursuit, fixation) and gaze patterns, making it an indispensable tool across multiple clinical domains (Uimonen et al., 2024; Zibold et al., 2024).

Additionally, VR systems can incorporate sensors such as pressure and motion detectors, as well as electrodes that measure electrical fields, skin conductance, or temperature. These integrations allow healthcare providers to efficiently monitor patient states and track progress in real time. For example, in the field of physical therapy, VR has been utilized to gamify rehabilitation exercises, thereby enhancing patient engagement and promoting active participation in their therapeutic processes. This approach not only increases patient adherence but also facilitates the objective measurement of performance parameters, such as range of motion or motor coordination (Laver et al., 2017).

In psychology and mental health, VR has found significant clinical applications, including the treatment of pain, stress, and anxiety. By immersing patients in virtual environments designed to distract or calm them, VR can effectively alleviate psychological symptoms by shifting focus away from real-world stressors (Malloy & Milling, 2010) On the other hand, VR also serves as a powerful tool in exposure therapy, allowing patients to confront and process phobias or traumatic memories in a controlled and safe virtual setting. This method has been shown to increase the patient’s anxiety threshold and reduce sensitivities, offering an alternative treatment option when traditional exposure techniques are hindered by logistical challenges or patient reluctance (Carl et al., 2019; Javvaji et al., 2024).

Like many other areas of healthcare, ophthalmology faces significant challenges. Access to eye care, including visits to ophthalmologists and vision screenings, is a pressing public health concern, particularly for individuals in remote or underserved areas. A considerable number of visual impairments go undiagnosed each year due to insufficient screenings and underdiagnosis (Mergen & Ramsey, 2021). Socioeconomic disparities remain a major barrier to accessing eye care, as individuals from lower socioeconomic backgrounds and those without private insurance are less likely to engage in routine eye screenings (Lou et al., 2017; Zhang et al., 2012). In rural and remote communities, access to specialized ophthalmological services is particularly limited, even though teleophthalmology has alleviated some barriers by enabling remote consultations and screenings. However, physical access to specialized care continues to be a significant challenge in these regions. Additionally, immigrants and minority populations often encounter cultural and linguistic obstacles that further hinder their access to eye care services, highlighting the need for culturally sensitive health communication strategies to improve participation in vision screening programs (Sekimitsu et al., 2025).

Moreover, traditional in-office diagnostic procedures, such as standard automated perimetry, can be both costly and uncomfortable for patients, which may deter individuals from seeking necessary care or following through with recommended screenings. Studies have shown that automated perimetry, while essential for diagnosing and monitoring conditions like glaucoma, can present financial burdens (Chauhan et al., 2008; Traverso et al., 2005), and the testing procedure itself can be inconvenient for a significant proportion of patients (Hicks et al., 2023). These factors, including cost and perceived discomfort, have been identified as barriers to glaucoma care and follow-up, potentially leading to delays in diagnosis or treatment (Myint et al., 2010; Newman-Casey et al., 2015). Another key limitation of existing ophthalmic tests is their lack of specificity and accuracy. Many traditional tests focus solely on one eye at a time, potentially missing discrepancies between the eyes (Sunness et al., 1997) or failing to capture comprehensive visual function. Furthermore, existing tests typically do not integrate auditory stimuli, which can be crucial for assessing multisensory integration and spatial awareness (Lewald & Guski, 2003). These limitations highlight the need for more precise and holistic diagnostic tools in ophthalmology.

Virtual Reality (VR) tools have emerged as promising technologies for the future of vision care, demonstrating significant utility in the screening, diagnosis, and treatment of various eye diseases.

### Screening in Vision Care

In the context of vision screening, VR offers several advantages over traditional methods, particularly by providing an immersive environment that enhances patient engagement and reduces the impact of external distractions. VR-based screening tools are gaining attention as a potential solution for improving accessibility to vision tests, especially in underserved or remote areas. These systems can facilitate comprehensive screenings by integrating various visual stimuli, including those for visual field testing, contrast sensitivity, and peripheral vision assessments. The ability of VR to offer a more controlled and standardized testing environment, with precise control over light conditions and visual distractions, is a significant advantage over traditional methods (Groth et al., 2023; Tsapakis et al., 2017). Moreover, VR-based tools can provide immediate feedback to patients and clinicians, allowing for real-time adjustments and more personalized screening experiences (Wroblewski et al., 2014). Research has shown that VR can provide reliable results comparable to traditional vision screening techniques such as Humphrey Field Analyzer and Octopus 900, making it a valuable tool for early detection of conditions such as glaucoma and other retinal diseases (Phu et al., 2024; Stapelfeldt et al., 2021).

### VR for Diagnosis

Building on its potential for screening, VR has shown considerable promise in diagnosis as well. The immersive nature of VR headsets allows for precise simulations of visual field deficits, such as those seen in glaucoma, and can be used to track the progression of visual impairments over time. Studies have demonstrated that VR-derived visual field measurements are highly correlated with those obtained using conventional diagnostic tools like the Octopus 900 (Stapelfeldt et al., 2021), suggesting that VR could serve as an effective and cost-efficient diagnostic alternative (Razeghinejad et al., 2021; Stapelfeldt et al., 2021).

### VR for Treatment

In addition to its diagnostic applications, VR has also been explored as a treatment modality for various visual impairments (Cinnera et al., 2022; Daibert-Nido et al., 2021; Molina-Martín et al., 2023). By creating customized virtual environments for vision training, VR can help individuals improve visual acuity, depth perception, and eye-hand coordination. For instance, dichoptic VR training has been shown to significantly improve visual acuity and stereo acuity in patients with anisometropic amblyopia (Žiak et al., 2017). Furthermore, VR has shown promise in rehabilitation programs for conditions like hemianopia, offering patients a safe and engaging way to engage in therapy from home (Daibert-Nido et al., 2021). These findings suggest that VR has potential not only as a diagnostic tool but also as a versatile platform for personalized vision therapy and rehabilitation.

Despite a growing body of research highlighting the potential of virtual reality (VR) in ophthalmology, to our knowledge, no comprehensive review has yet synthesized the full scope of its applications across screening, diagnostic, and therapeutic domains, particularly in relation to the diverse research designs that have explored these areas.

### Objectives

This rapid review aims to synthesize the current state of peer-reviewed research on VR-based applications in vision care. Specifically, we address the following key aspects:

***A) Application Types*** – A comparison of VR applications used for screening, diagnosis, and intervention, with a focus on their targeted outcomes, reported validity and/or effectiveness, and the administration protocols (e.g., who administers the interventions, in what settings, duration/frequency of sessions, and integration with existing ophthalmological tools).

***B) Technical Properties*** – An examination of the technical specifications of the VR systems employed, including hardware (e.g., devices used) and software/content (e.g., the virtual environments or tasks utilized).

***C) Study Characteristics*** – An evaluation of study characteristics, including sample size, participant conditions, and the risk of bias, to assess the quality and generalizability of the findings.

## Methods

This review was conducted according to the PRISMA (Preferred Reporting Items for Systematic Reviews and Meta-Analyses) guidelines. The review protocol was registered in PROSPERO (International Prospective Register for Systematic Reviews) with the registration number <u>CRD42023456214</u>.

### Search Strategy

A comprehensive search strategy was developed using a combination of database-specific subject headings and text words for the main concepts of virtual reality and select eye diseases and auditory phenomenon. Results were limited to humans and English. Conference materials were excluded from Embase.

The databases Ovid MEDLINE, Ovid Embase, Cochrane Database of Systematic Reviews (Ovid), and Cochrane Central Register of Controlled Trials (Ovid) were searched from inception to August 14, 2023. Full search strategies are reported in <u>Appendix 1.</u> Additional relevant studies were included by searching the reference lists of included publications.

### Study selection

All relevant articles were inputted into Rayyan QCRI. To filter articles that focused on virtual reality and visual and auditory outcomes, a dual filtration system was used to isolate articles that included 1) ‘virtual reality’ or ‘VR’ and 2) ‘vision’, ‘visual’, ‘audition’, ‘audiology’ in the abstract or title. Duplications were detected and resolved on Rayyan; if the similarity percentage was less than 98%, additional reference screening was conducted to ensure the articles were duplicates. Secondary literature was excluded using the keywords ‘meta-analysis’, ‘review’, and ‘systematic review’.

### Inclusion and exclusion criteria

All primary research studies published in English and in peer-reviewed journals from up until the search date were included. There were no demographic restrictions. Studies were included if they included immersive and portable VR devices, namely head-mounted displays (HMDs), as the primary intervention. Our criteria for immersiveness is based on, factors including, display resolution, field of view, movement degrees of freedom, number of senses stimulated (hearing, vision, touch, and proprioception), the ability to track and update user inputs, and the ability to isolate the user from stimuli in the real world. Studies reporting outcomes that demonstrate the clinical effectiveness of VR as a therapeutic/diagnostic/screening tool for visual cognition or vision impairments with audition were included.

Studies were excluded if they were: 1) conducted with animals; 2) exclusively focused on non-immersive and not portable VR devices, such as 3D glasses and cave-automatic virtual environments (CAVEs); 3) had missing data within the study or outcomes not pertaining to our clinical area of interest (ie: vision or vision and audition); 4) secondary literature, such as reviews, meta-analyses, conference abstracts, commentaries, editorials, and full texts with no original data; or 5) full texts not accessible through the institutional library systems used within the process, including the ones provided by University Health Network, York University, McMaster University, and the University of Toronto.

The screening of title and abstract first involved two reviewers independently screening 20 abstracts to assess inter-rater reliability. Discrepancies were resolved by consensus, and the inclusion criteria were further clarified to include studies pertaining to visual cognition or perception. The remaining abstracts were then divided in half and screened separately. Each reviewer conducted randomized cross-checks of each other’s articles to ensure agreement and reliability of screening.

### Data extraction (coding and categorizing studies)

A comprehensive full-text evaluation was performed by two reviewers independently. Qualitative and quantitative data from each article were coded and categorized into the following aspects:

1) research type and theme; 2) purpose of intervention; 3) study demographics; 4) novelty and attributes of VR tool. Additional information collected and the full coding spreadsheet are provided in <u>Appendix 2</u>.

The conceptualization of terms was discussed during the early stages of coding and was further cross-checked by both reviewers to minimize subjective differences. For example, therapeutic interventions are defined as tools that are used to improve an existing condition, screening interventions as tools to detect whether participants have a certain condition, and diagnostic interventions as tools to confirm the condition. <u>Appendix 3</u> provides the full definition of concepts used for the full-text screening of articles. After confirming agreement on conceptual definitions, the reviewers reverse-coded each other’s articles.

### Quality assessment

Two reviewers independently assessed the quality of each eligible study using the Mixed Methods Appraisal Tool (MMAT, Version 2018). The MMAT was chosen as it allows the critical appraisal of the methodological quality of five major research designs: qualitative research, randomized controlled trials, non-randomized studies, quantitative descriptive studies, and mixed methods studies. Each study was analyzed manually using the MMAT checklist. After selecting the most appropriate study category to appraise, responses were noted as ‘yes’, ‘no’, or ‘unclear’ according to the respective methodological criteria.

To further inform analysis, studies were coded according to the Virtual Reality Clinical Outcomes Research Experts (VR-CORE) model as VR1, VR2, or VR3 studies (Birckhead et al., 2019). VR1 studies focused on content development, VR2 studies focused on the early testing of feasibility and initial clinical efficacy, and VR3 studies were RCTs comparing clinical outcomes between intervention and control groups.

#### Analysis

After categorizing each publication based on several characteristics, descriptive statistics, specifically the frequency of publications allocated within each category, were determined. Additionally, the proportion of studies within each sub-category was computed, whereby each frequency was divided by the total number of publications encompassed in the analysis.

## Results

As outlined in the PRISMA diagram (Figure 1), 7,324 citations were initially collected from the specified databases and processed using Rayyan QCRI. After duplicate removal and further filtration, 1,246 citations proceeded to the first screening stage, where secondary reviews were excluded. Following the exclusion of 69 review articles, 1,177 articles were screened at the abstract stage, resulting in the exclusion of 977 articles. A total of 196 articles underwent full-text screening, with 120 being excluded due to failure to meet one or more eligibility criteria. Ultimately, 76 publications were included for analysis (<u>Appendix 4</u>).

**Figure 1:**
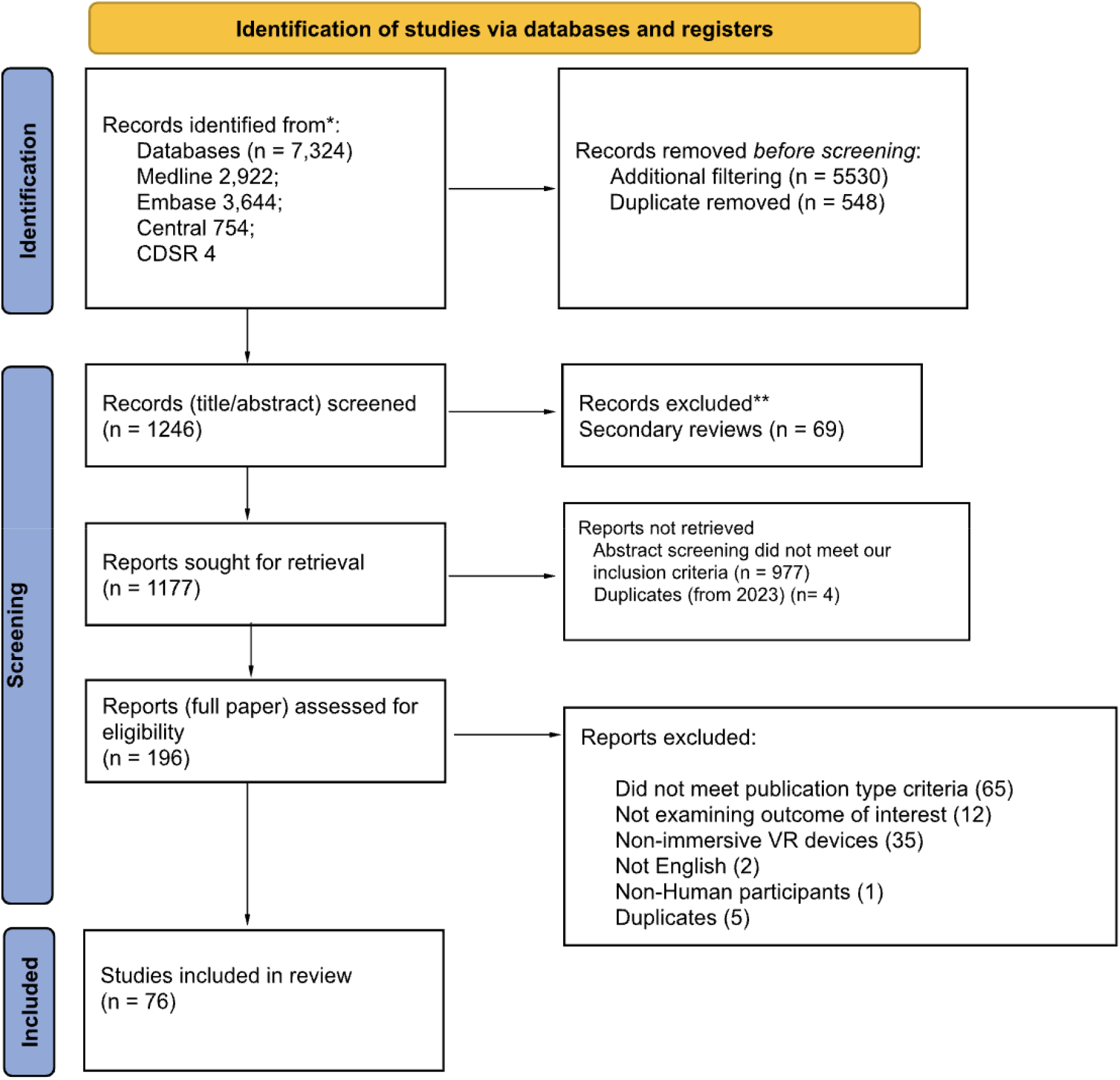
**PRISMA Diagram** depicting the filtration process from initial retrieval to final inclusion in the qualitative synthesis.

The studies were categorized according to the VR-CORE model. Sixty-four studies (84.2%) were classified as VR2, which include non-randomized research exploring the effectiveness of VR interventions in vision care. In contrast, twelve studies (15.8%) were classified as VR3, representing randomized-controlled trials (RCTs). Table 1 provides a breakdown of the study characteristics, highlighting the distribution of research designs and interventions across the 76 publications.

**Table 1:** Overview of study characteristics.

| Research Type |  | Research Theme |  | Intervention Use |  |
| --- | --- | --- | --- | --- | --- |
| Comparator Interventions | 25 (32.9%) | Visual Impairment | 58 (76.3%) | Therapeutic | 31 (40.8%) |
| Outcomes: validated measures | 22 (28.9%) | Visual Cognition | 16 (21.1%) | Diagnostic | 29 (38.2%) |
| RCTs | 12 (15.8%) | Visual Impairment + Audition | 1 (1.3%) | Screening | 16 (21.0%) |
| Pilot Trials | 12 (15.8%) | Visual Cognition + Audition | 1 (1.3%) |  |  |
| Case Report | 4 (5.3%) |  |  |  |  |
| Cross-sectional | 1 (1.3%) |  |  |  |  |

Additionally, the nature of VR interventions was analyzed to determine whether they utilized novel or existing methodologies. Of the studies reviewed, 62 (81.6%) introduced novel approaches, while 14 (18.4%) incorporated existing methodologies into VR systems. For example, the study by Chen et al. (2022) employed a novel software (LUXIE) for visual field testing integrated into VR headsets, while Kim et al. (2019) implemented the established King-Devick Test Chart within a VR setup.

Table 2 offers an overview of participant demographics, including sample sizes and distributions across various conditions. A total of 11 medical conditions were classified as “other,” encompassing general visual impairments, retinitis pigmentosa, low vision, inherited retinal degeneration, congenital visual impairments, and ptosis. Notably, some studies simulated the effects of eye diseases on the field of vision in healthy participants. For instance, Neugebauer et al. (2021) simulated retinitis pigmentosa, and Massiceti et al. (2018) simulated low vision and blindness. These methodologies carry limitations, as healthy participants do not experience real-world visual field loss or develop compensatory strategies naturally.

**Table 2:** Overview of study Participant characteristics.

| Participant Age Group |  | Participant Medical Condition |  | Gross sample size |  |
| --- | --- | --- | --- | --- | --- |
| Patients - Adult | 42 (55.3%) | Glaucoma | 20 (26.3%) | 1-19 | 29 (38.2%) |
| Healthy - Adult | 13 (17.1%) | Healthy | 18 (23.7%) | 20-49 | 24 (31.6%) |
| Patients - Pediatric | 9 (11.8%) | Amblyopia and Other Binocular Vision Disorders | 15 (19.7%) | 50-99 | 14 (18.4%) |
| Patients - Adult & Pediatric | 8 (10.5%) | Neuro-Ophthalmic Disorder | 7 (9.2%) | 100-199 | 7 (9.2%) |
| Healthy - Pediatric | 4 (5.3%) | Stroke and Visual Neglect | 5 (6.6%) | 200+ | 2 (2.6%) |
|  |  | Other | 11 (14.5%) |  |  |

Table 3 presents a summary of the head-mounted display (HMD) brands and models used in the 76 studies. The analysis revealed a diverse range of HMD devices, with some devices appearing in only one study (categorized as “other”). Popular models include Oculus Rift, Fove 0, Oculus Quest, and Oculus Go.

**Table 3:** Overview of HMD brands and models.

| HMD Device Brand/Model |  |
| --- | --- |
| HTC ViVE | 14 (18.4%) |
| Oculus Rift | 11 (14.5%) |
| Oculus Go | 7 (9.2%) |
| Olleyes VisuALL | 6 (7.9%) |
| Unspecified HMD | 5 (6.6%) |
| VR OnePlus | 4 (5.3%) |
| Oculus Quest | 3 (4.0%) |
| Pico neo | 3 (4.0%) |
| C3 Visual Field Analyzer | 2 (2.6%) |
| Fove 0 | 2 (2.6%) |
| Samsung Gear | 2 (2.6%) |
| VR Shinecon | 2 (2.6%) |
| Other | 15 (19.7%) |

Regarding the VR system setup, 39 studies (51.3%) utilized tethered HMDs, while 36 studies (47.4%) employed stand-alone devices that did not require external hardware. Only one study (1.3%) did not specify the HMD device used.

Industry collaboration was another key aspect analyzed. Twelve studies (15.8%) were identified as joint ventures with VR companies, while 43 studies (56.6%) had unclear or unspecified levels of collaboration. A total of 21 studies (27.6%) were developed without industry involvement.

Regarding the journals in which these studies were published, 47 (61.8%) appeared in ophthalmology-focused journals, 21 (27.6%) in general health journals, and 5 (6.6%) in technology-oriented journals. Three publications (4.0%) were categorized as “other” and appeared in journals such as Cyberpsychology, *Behavior and Social Networking, Military Medicine*, and the *American Journal of Case Reports*.

Each publication was evaluated using the MMAT tool to assess study quality and risk of bias. The results are provided in Appendix 2.

## Discussion

The advancement of VR technology has opened new possibilities in ophthalmology, particularly in the screening and diagnosis of eye and visual diseases. However, as demonstrated in our study, the application of VR in vision care remains in its early stages and does not come without challenges.

One of the primary obstacles in integrating VR headsets into ophthalmological practices, as suggested by our study, is the need for standardization and validation. The results from our review reveal a notable variation in study designs and sample sizes, which further complicates the process of establishing consistent and reliable VR-based diagnostic tools. Specifically, 64 studies (84.2%) were classified as VR2, representing non-randomized research exploring the effectiveness of VR interventions, while only 12 studies (15.8%) were randomized-controlled trials (VR3). This disparity highlights the need for more rigorous, randomized clinical trials to establish the efficacy and reliability of VR technologies in diagnosing and monitoring eye diseases.

Additionally, the variation in population characteristics across the studies adds another layer of complexity. While some studies focused on specific visual impairments such as retinitis pigmentosa, amblyopia, or glaucoma, others simulated eye conditions in healthy participants. These differences in study populations emphasize the necessity for standardized protocols and clear guidelines for the application of VR in vision care. The diverse sample sizes and conditions examined in the reviewed studies underscore the lack of a unified approach in the field, making it difficult to draw broad conclusions about the generalizability and accuracy of VR diagnostics across different patient demographics.

Each VR system also presents its own set of specifications and performance metrics, further hindering the consistency of results. The varying devices, such as the Oculus Rift, Oculus Quest, and other head-mounted displays, differ in terms of resolution, field of view, and eye-tracking capabilities. As our study revealed, 51.3% of the studies employed tethered devices, while 47.4% used stand-alone systems. This diversity in VR hardware complicates the establishment of uniform standards and protocols for clinical use. As such, the future of VR in ophthalmology relies on the validation of these systems through rigorous comparative studies with traditional diagnostic tools, as well as the development of standardized protocols to ensure consistent performance across different devices and settings.

Ensuring that VR technology is user-friendly and accessible is critical for its successful implementation in ophthalmology. Our study highlights the diversity in the types of VR devices used across studies, with 51.3% of the studies utilizing tethered headsets (e.g., Oculus Rift, Fove 0) and 47.4% employing standalone systems (e.g., Oculus Quest, Oculus Go). This distinction is significant in terms of usability, as tethered systems often require additional equipment such as computers or external sensors, which may complicate their use in clinical settings. On the other hand, standalone devices, while potentially more user-friendly due to their portability and ease of setup, may face limitations in terms of processing power or battery life. These differences underline the need to consider the specific requirements of both healthcare providers and patients when selecting VR equipment for clinical or home-based use.

Many patients, particularly elderly individuals or those with limited technological proficiency, may find it challenging to use VR headsets. To overcome this barrier, it is essential to design intuitive interfaces and provide comprehensive training for both patients and healthcare providers. Furthermore, addressing issues such as comfort, ease of use, and potential side effects like motion sickness or eye strain is crucial for ensuring patient compliance and satisfaction. As noted in several studies, including those in our review, the use of VR technology in clinical settings often involves a learning curve, and patients may experience discomfort if the headsets are not well-fitted or if the user interface is overly complex (Astek et al., 2024; Kim & Shin, 2020).

In addition to usability concerns, the use of VR headsets for medical purposes necessitates the collection and processing of sensitive patient data. This introduces privacy and security risks, which were highlighted in several of the studies we reviewed. Ensuring compliance with privacy regulations such as HIPAA in the United States or GDPR in Europe is critical to safeguard patient information. Our study highlights the importance of navigating the regulatory landscape, with 15.8% of the publications identified as being jointly developed in collaboration with industry partners. These collaborations may help accelerate the development of VR systems by providing access to cutting-edge technologies and expertise, but they also raise concerns about commercialization and the potential for conflicts of interest. The implementation of robust cybersecurity measures, including data encryption, secure storage, and regular security audits, is equally essential. Clear policies and protocols must be established to govern data access and sharing, ensuring that patient data is used ethically and responsibly while maintaining trust in the system (Bull et al., 2015). Moreover, ethical considerations such as informed consent, the potential for over-reliance on technology, and the impact on patient-provider relationships must be carefully managed to maintain trust and integrity in healthcare delivery. This includes understanding the potential biases introduced by industry collaborations and ensuring that VR-based interventions remain patient-centered.

Importantly, VR headsets must meet high technical standards to be effective in medical applications. As noted in several studies, issues such as resolution, field of view, luminance range, and tracking accuracy can affect the reliability of VR-based diagnostics. For instance, Stapelfeldt et al. (2021) highlighted that limited luminance range in their VR system prevented effective analysis of patients with advanced glaucoma. This issue, along with concerns regarding battery life, durability, and compatibility with other medical devices, underscores the need for VR headsets to meet high technical standards in order to be effective in medical applications. Future developments in VR for ophthalmology must prioritize these technical aspects to ensure that the technology can be used reliably for diagnostic and therapeutic purposes. The integration of artificial intelligence (AI) with VR technology has the potential to significantly enhance diagnostic accuracy and efficiency. AI algorithms can process data from VR systems to identify early signs of diseases such as glaucoma and macular degeneration. Additionally, VR headsets offer the advantage of continuous monitoring, which allows for ongoing assessment of patients’ visual health. This real-time data collection can help create more personalized treatment plans and prompt interventions when needed. As VR technology becomes more affordable, it could emerge as a cost-effective solution for large-scale vision screenings, alleviating the burden on healthcare systems and improving access to eye care services on a broader scale.

Finally, while VR technology has the potential to be a cost-effective solution in the long run, the initial investment in VR headsets and related infrastructure can be significant. Healthcare providers must consider the cost of purchasing, maintaining, and updating VR equipment. Furthermore, there may be additional costs associated with training staff and integrating VR systems into existing healthcare workflows. Securing funding and demonstrating the cost-benefit ratio of VR technology in terms of improved patient outcomes and operational efficiency will be crucial for its adoption.

### Limitations

While this rapid review followed established protocols to ensure methodological rigor, several limitations should be noted. First, the exclusion of conference materials from Embase may have resulted in the absence of relevant studies that were not yet fully peer-reviewed or were presented at conferences. Although the inclusion criteria were comprehensive, limiting studies to those published in English potentially excluded valuable research published in other languages. Additionally, while we aimed to capture all relevant studies, our search strategy was restricted to four databases, which might not have encompassed all existing research on virtual reality (VR) in vision care, especially in grey literature or unpublished works. The dual filtration system for study selection was robust; however, there could have been inconsistencies in the interpretation of inclusion/exclusion criteria despite rigorous training and random cross-checks between reviewers. Moreover, the process of data extraction was highly dependent on the quality and completeness of the studies included, missing or unclear data in some articles could have introduced bias or limited the depth of our analysis. The use of the MMAT tool for quality assessment, while appropriate, may be subject to reviewer interpretation, and we acknowledge that some studies may not have been appraised with the same level of scrutiny across all categories. Finally, our approach to categorizing studies according to the VR-CORE model and allocating studies into VR1, VR2, or VR3 categories was based on a conceptual framework that might not fully capture the nuanced differences in study designs or interventions.

## Conclusion and Future Directions

In conclusion, VR holds great promise for transforming the field of ophthalmology by improving the accessibility, accuracy, and efficiency of vision care. Although VR-based applications for vision screening, diagnosis, and treatment are still in the early stages, the potential for VR to revolutionize eye care is evident. However, to fully realize its benefits, further research is required to establish its clinical efficacy, standardized protocols, and refine technical aspects of the technology. Additionally, efforts to improve the user-friendliness, affordability, and ethical considerations of VR will be essential for ensuring its successful integration into healthcare systems. Moving forward, continued collaboration between healthcare providers, researchers, and technology developers will be vital to advancing VR-based solutions in vision care.

## Supporting information

Search strategy and terms

Coding MMAT

Definition of codes

List of reviewed articles

## Data Availability

All data produced in the present study are available upon reasonable request to the authors

## Availability of data and materials

All data generated or analysed during this study are included in this published article and its supplementary information files.

## Authors’ contributions

## Acknowledgements

None.

