## Supplementary material for "Seeing Opportunity in Virtual Reality: A Rapid Review of the Use of VR as a Tool in Vision Care": Search strategy and terms

### AppSearch Methods for Virtual Reality and Eye Diseases

#### Search Methods

A comprehensive search strategy was developed using a combination of database-specific subject headings and text words for the main concepts of virtual reality and select eye diseases and auditory phenomenon. Results were limited to human and English. Conference materials were excluded from Embase.

We searched the following databases on August 14, 2023 and August 16, 2024: Ovid MEDLINE; Ovid Embase; Cochrane Database of Systematic Reviews (Ovid); and Cochrane Central Register of Controlled Trials (Ovid). Full search strategies in Appendix I.

The reference lists of included publications were also searched and considered for inclusion.

#### Appendix I

Searches as run on August 14, 2023

Ovid MEDLINE(R) ALL <1946 to August 11, 2023>

| **#** | **Searches** | **Results** | **Type** |
| --- | --- | --- | --- |
| 1 | exp eye diseases/ | 638261 | Advanced |
| 2 | Optic Atrophy/ge | 757 | Advanced |
| 3 | exp Optic Nerve/ab | 1994 | Advanced |
| 4 | Visual Fields/ | 32529 | Advanced |
| 5 | exp Vision Tests/ | 117742 | Advanced |
| 6 | exp Auditory Perception/ | 85151 | Advanced |
| 7 | Sound Localization/ | 4818 | Advanced |
| 8 | (eye adj2 disease*).mp. | 54429 | Advanced |
| 9 | (eye adj2 disorder*).mp. | 2214 | Advanced |
| 10 | Albinism.mp. | 4426 | Advanced |
| 11 | (Chediak adj2 higashi syndrome).mp. | 1210 | Advanced |
| 12 | (Congenital adj2 amauros?s).mp. | 1569 | Advanced |
| 13 | dysgenesis neuroepithelialis retinae.mp. | 1 | Advanced |
| 14 | hereditary epithelial dysplasia of retina.mp. | 0 | Advanced |
| 15 | hereditary retinal aplasia.mp. | 0 | Advanced |
| 16 | heredoretinopathia congenitalis.mp. | 0 | Advanced |
| 17 | leber abiotroph*.mp. | 0 | Advanced |
| 18 | (leber* adj2 amauros?s).mp. | 1578 | Advanced |
| 19 | leber congenital tapetoretinal degeneration.mp. | 0 | Advanced |
| 20 | hereditary optic atroph*.mp. | 127 | Advanced |
| 21 | (optic adj2 hypoplasia*).mp. | 837 | Advanced |
| 22 | (retinal adj2 degeneration*).mp. | 16737 | Advanced |
| 23 | (retinal adj2 dysplasia*).mp. | 488 | Advanced |
| 24 | pigmentary retinopathy*.mp. | 612 | Advanced |
| 25 | retinitis pigmentosa.mp. | 12474 | Advanced |
| 26 | tapetoretinal degeneration*.mp. | 218 | Advanced |
| 27 | fundus flavimaculatus.mp. | 231 | Advanced |
| 28 | macular dystrophy with flecks.mp. | 2 | Advanced |
| 29 | stargardt disease.mp. | 1072 | Advanced |
| 30 | cod md syndrome*.mp. | 2 | Advanced |
| 31 | cerebromuscular dystrophy.mp. | 12 | Advanced |
| 32 | cerebroocular dysplasia muscular dystrophy syndrome.mp. | 0 | Advanced |
| 33 | chemke syndrome.mp. | 1 | Advanced |
| 34 | congenital muscular dystrophy dystroglycanopathy.mp. | 6 | Advanced |
| 35 | walker-warburg syndrome*.mp. | 471 | Advanced |
| 36 | fukuyama cmd.mp. | 27 | Advanced |
| 37 | (fukuyama adj3 muscular dystrophy).mp. | 410 | Advanced |
| 38 | fukuyama syndrome.mp. | 1 | Advanced |
| 39 | hard syndrome*.mp. | 4 | Advanced |
| 40 | lgmd2k.mp. | 6 | Advanced |
| 41 | mddga1.mp. | 0 | Advanced |
| 42 | meb syndrome.mp. | 0 | Advanced |
| 43 | muscle-eye-brain syndrome.mp. | 3 | Advanced |
| 44 | muscle eye brain disease*.mp. | 190 | Advanced |
| 45 | muscular dystrophy due to defective glycosylation of dystroglycan 4a.mp. | 0 | Advanced |
| 46 | muscular dystrophy-dystroglycanopathy.mp. | 30 | Advanced |
| 47 | pagon syndrome*.mp. | 0 | Advanced |
| 48 | warburg syndrome.mp. | 488 | Advanced |
| 49 | alpha dystroglycanopathies.mp. | 48 | Advanced |
| 50 | congenital mesodermal dysmorphodystroph*.mp. | 0 | Advanced |
| 51 | gems.mp. | 1235 | Advanced |
| 52 | glaucoma-lens ectopia-microspherophakia-stiffness-shortness syndrome.mp. | 1 | Advanced |
| 53 | (marchesani adj2 syndrome*).mp. | 209 | Advanced |
| 54 | spherophakia brachymorphia syndrome*.mp. | 5 | Advanced |
| 55 | cataract*.mp. | 78249 | Advanced |
| 56 | lens opacity*.mp. | 1093 | Advanced |
| 57 | pseudoaphakia*.mp. | 7 | Advanced |
| 58 | (lens adj2 cloud*).mp. | 60 | Advanced |
| 59 | glaucoma*.mp. | 82175 | Advanced |
| 60 | increased intraocular pressure.mp. | 1449 | Advanced |
| 61 | ocular hypertension.mp. | 9884 | Advanced |
| 62 | optic neuropathy.mp. | 11709 | Advanced |
| 63 | brown* tendon sheath syndrome*.mp. | 4 | Advanced |
| 64 | conjugate gaze spasm*.mp. | 0 | Advanced |
| 65 | convergence excess*.mp. | 114 | Advanced |
| 66 | convergence insufficienc*.mp. | 560 | Advanced |
| 67 | cyclophoria*.mp. | 24 | Advanced |
| 68 | eye motility disorder*.mp. | 18 | Advanced |
| 69 | eye movement disorder*.mp. | 452 | Advanced |
| 70 | internuclear ophthalmoplegia*.mp. | 722 | Advanced |
| 71 | ocular motility disorder*.mp. | 4435 | Advanced |
| 72 | ocular torticollis.mp. | 116 | Advanced |
| 73 | opsoclonus.mp. | 1116 | Advanced |
| 74 | parinaud* syndrome*.mp. | 333 | Advanced |
| 75 | paroxysmal ocular dyskinesia*.mp. | 0 | Advanced |
| 76 | pseudoophthalmoplegia*.mp. | 0 | Advanced |
| 77 | skew deviation*.mp. | 327 | Advanced |
| 78 | smooth pursuit deficienc*.mp. | 2 | Advanced |
| 79 | spasm of conjugate gaze.mp. | 0 | Advanced |
| 80 | tendon sheath syndrome of brown.mp. | 5 | Advanced |
| 81 | fisher syndrome.mp. | 1348 | Advanced |
| 82 | miller fisher.mp. | 1343 | Advanced |
| 83 | (ophthalmoplegia, ataxia and areflexia syndrome).mp. | 5 | Advanced |
| 84 | nystagmus.mp. | 19408 | Advanced |
| 85 | involuntary eye movement*.mp. | 164 | Advanced |
| 86 | Jerky eye movement*.mp. | 11 | Advanced |
| 87 | cranial nerve iii disease*.mp. | 0 | Advanced |
| 88 | oculomotor nerve disease*.mp. | 1845 | Advanced |
| 89 | oculomotor nerve disorder*.mp. | 2 | Advanced |
| 90 | oculomotor nerve pals*.mp. | 977 | Advanced |
| 91 | oculomotor nerve paralys?s.mp. | 77 | Advanced |
| 92 | oculomotor neuropath*.mp. | 25 | Advanced |
| 93 | third cranial nerve disease*.mp. | 0 | Advanced |
| 94 | third nerve pals*.mp. | 815 | Advanced |
| 95 | third nerve paralysis.mp. | 39 | Advanced |
| 96 | ophthalmoplegia*.mp. | 12334 | Advanced |
| 97 | oculomotor paralysis.mp. | 1024 | Advanced |
| 98 | ophthalmopares?s.mp. | 552 | Advanced |
| 99 | dancing eyes.mp. | 41 | Advanced |
| 100 | myoclonic encephalopathy*.mp. | 270 | Advanced |
| 101 | kinsbourne syndrome.mp. | 21 | Advanced |
| 102 | opsoclonus myoclonus.mp. | 778 | Advanced |
| 103 | strabismus.mp. | 20277 | Advanced |
| 104 | dissociated horizontal deviation*.mp. | 27 | Advanced |
| 105 | dissociated vertical deviation*.mp. | 305 | Advanced |
| 106 | heterophoria*.mp. | 554 | Advanced |
| 107 | heterotropia*.mp. | 136 | Advanced |
| 108 | hypertropia*.mp. | 519 | Advanced |
| 109 | phoria*.mp. | 611 | Advanced |
| 110 | squint*.mp. | 2041 | Advanced |
| 111 | crossed eye*.mp. | 51 | Advanced |
| 112 | tolosa hunt syndrome*.mp. | 579 | Advanced |
| 113 | Ametropia*.mp. | 1080 | Advanced |
| 114 | refractive disorder*.mp. | 85 | Advanced |
| 115 | refractive error*.mp. | 16494 | Advanced |
| 116 | aniseikonia.mp. | 559 | Advanced |
| 117 | anisometropia.mp. | 2231 | Advanced |
| 118 | astigmatism*.mp. | 14185 | Advanced |
| 119 | corneal wavefront aberration*.mp. | 989 | Advanced |
| 120 | farsighted*.mp. | 141 | Advanced |
| 121 | longsighted*.mp. | 3 | Advanced |
| 122 | hypermetropia*.mp. | 739 | Advanced |
| 123 | hyperopia*.mp. | 5457 | Advanced |
| 124 | myopia*.mp. | 28444 | Advanced |
| 125 | nearsighted*.mp. | 168 | Advanced |
| 126 | shortsighted*.mp. | 151 | Advanced |
| 127 | presbyopia*.mp. | 2625 | Advanced |
| 128 | (retinal adj2 disease*).mp. | 31618 | Advanced |
| 129 | (cone adj2 dystroph*).mp. | 1661 | Advanced |
| 130 | diabetic retinopath*.mp. | 40677 | Advanced |
| 131 | hypertensive retinopath*.mp. | 897 | Advanced |
| 132 | diabetic eye disease*.mp. | 538 | Advanced |
| 133 | diabetic macular edema.mp. | 4752 | Advanced |
| 134 | blindness.mp. | 48522 | Advanced |
| 135 | hemeralopia*.mp. | 139 | Advanced |
| 136 | macropsia*.mp. | 57 | Advanced |
| 137 | metamorphopsia*.mp. | 979 | Advanced |
| 138 | micropsia*.mp. | 109 | Advanced |
| 139 | vision disabilit*.mp. | 77 | Advanced |
| 140 | vision disorder*.mp. | 31032 | Advanced |
| 141 | vision impairment*.mp. | 2905 | Advanced |
| 142 | visual disorder*.mp. | 1084 | Advanced |
| 143 | visual impairment*.mp. | 15200 | Advanced |
| 144 | visual disabilit*.mp. | 1062 | Advanced |
| 145 | amblyopia*.mp. | 10382 | Advanced |
| 146 | lazy eye*.mp. | 66 | Advanced |
| 147 | amauros?s.mp. | 3525 | Advanced |
| 148 | sudden visual loss*.mp. | 289 | Advanced |
| 149 | achromatopsia*.mp. | 664 | Advanced |
| 150 | colo?r vision defect*.mp. | 4386 | Advanced |
| 151 | colo?r vision deficienc*.mp. | 534 | Advanced |
| 152 | colo?r vision impairment*.mp. | 89 | Advanced |
| 153 | colo?r perception deficienc*.mp. | 3 | Advanced |
| 154 | deutan defect.mp. | 7 | Advanced |
| 155 | monochromatopsia.mp. | 0 | Advanced |
| 156 | protan defect.mp. | 7 | Advanced |
| 157 | tritan defect.mp. | 22 | Advanced |
| 158 | diplopia*.mp. | 12796 | Advanced |
| 159 | double vision.mp. | 1396 | Advanced |
| 160 | polyopsia*.mp. | 3 | Advanced |
| 161 | nyctalopia.mp. | 439 | Advanced |
| 162 | light sensitivit*.mp. | 2111 | Advanced |
| 163 | photophobia*.mp. | 4224 | Advanced |
| 164 | scotoma*.mp. | 5570 | Advanced |
| 165 | retinocochleocerebral vasculopath*.mp. | 16 | Advanced |
| 166 | susac* syndrome.mp. | 474 | Advanced |
| 167 | diminished vision.mp. | 150 | Advanced |
| 168 | reduced vision.mp. | 883 | Advanced |
| 169 | subnormal vision.mp. | 97 | Advanced |
| 170 | sub-normal vision.mp. | 9 | Advanced |
| 171 | low vision.mp. | 3451 | Advanced |
| 172 | macular degeneration*.mp. | 31253 | Advanced |
| 173 | maculopath*.mp. | 5645 | Advanced |
| 174 | macular dystroph*.mp. | 1840 | Advanced |
| 175 | macular disorder*.mp. | 182 | Advanced |
| 176 | retinal degeneration*.mp. | 15800 | Advanced |
| 177 | retinal degenerative disease*.mp. | 1143 | Advanced |
| 178 | visual field*.mp. | 52013 | Advanced |
| 179 | vision test*.mp. | 12007 | Advanced |
| 180 | colo?r perception test*.mp. | 2609 | Advanced |
| 181 | ocular refraction*.mp. | 242 | Advanced |
| 182 | vision screening.mp. | 3102 | Advanced |
| 183 | visual acuit*.mp. | 121036 | Advanced |
| 184 | visual contrast sensitivity*.mp. | 231 | Advanced |
| 185 | emmetropia*.mp. | 1905 | Advanced |
| 186 | automated perimetry exam*.mp. | 9 | Advanced |
| 187 | campimetr*.mp. | 313 | Advanced |
| 188 | perimetr*.mp. | 8234 | Advanced |
| 189 | tangent screen exam*.mp. | 13 | Advanced |
| 190 | auditory perception.mp. | 33112 | Advanced |
| 191 | auditory processing.mp. | 5136 | Advanced |
| 192 | (auditory adj2 localization*).mp. | 474 | Advanced |
| 193 | (sound adj2 localization*).mp. | 5866 | Advanced |
| 194 | spatial hearing.mp. | 527 | Advanced |
| 195 | or/1-194 | 882861 | Advanced |
| 196 | Virtual Reality/ | 5607 | Advanced |
| 197 | (virtual adj3 realit*).mp. | 18765 | Advanced |
| 198 | (augmented adj3 realit*).mp. | 4774 | Advanced |
| 199 | VRET.mp. | 136 | Advanced |
| 200 | iVR.mp. | 1929 | Advanced |
| 201 | iVRs.mp. | 210 | Advanced |
| 202 | iVE.mp. | 1881 | Advanced |
| 203 | iVEs.mp. | 180 | Advanced |
| 204 | (immers* adj2 environment*).mp. | 793 | Advanced |
| 205 | samsung gear.mp. | 40 | Advanced |
| 206 | oculus rift.mp. | 126 | Advanced |
| 207 | HCD vive.mp. | 0 | Advanced |
| 208 | hololens.mp. | 342 | Advanced |
| 209 | gear VR.mp. | 16 | Advanced |
| 210 | head-mount*.mp. | 2577 | Advanced |
| 211 | (head adj3 gear*).mp. | 97 | Advanced |
| 212 | HMD.mp. | 1370 | Advanced |
| 213 | Gen2-VR.mp. | 1 | Advanced |
| 214 | (google adj2 daydream).mp. | 4 | Advanced |
| 215 | (google adj2 cardboard).mp. | 24 | Advanced |
| 216 | PlayStation.mp. | 89 | Advanced |
| 217 | (ReTrak adj2 Utopia).mp. | 0 | Advanced |
| 218 | NOON VR.mp. | 0 | Advanced |
| 219 | Freefly VR.mp. | 0 | Advanced |
| 220 | Homido.mp. | 0 | Advanced |
| 221 | iWear.mp. | 0 | Advanced |
| 222 | (mixed adj2 realit*).mp. | 953 | Advanced |
| 223 | (simulat* adj2 environment*).mp. | 5866 | Advanced |
| 224 | (virtual adj2 environment*).mp. | 6199 | Advanced |
| 225 | Video Games/ | 7236 | Advanced |
| 226 | (computer adj2 game*).mp. | 1779 | Advanced |
| 227 | (video adj2 game*).mp. | 9697 | Advanced |
| 228 | gaming.mp. | 5541 | Advanced |
| 229 | user-computer interface/ | 39425 | Advanced |
| 230 | (user adj2 interface*).mp. | 47083 | Advanced |
| 231 | (virtual adj2 interface*).mp. | 194 | Advanced |
| 232 | Oculus Quest.mp. | 34 | Advanced |
| 233 | Meta Quest.mp. | 9 | Advanced |
| 234 | Metaverse.mp. | 313 | Advanced |
| 235 | (simulate* adj2 realit*).mp. | 108 | Advanced |
| 236 | (tethered adj2 headset*).mp. | 1 | Advanced |
| 237 | Serious game.mp. | 757 | Advanced |
| 238 | or/196-237 | 90536 | Advanced |
| 239 | 195 and 238 | 2875 | Advanced |
| 240 | animals/ not (animals/ and humans/) | 5111492 | Advanced |
| 241 | 239 not 240 | 2808 | Advanced |
| 242 | limit 241 to english language | 2714 | Advanced |
| 243 | remove duplicates from 242 | 2710 | Advanced |

Embase <1974 to 2023 August 11>

| **#** | **Searches** | **Results** | **Type** |
| --- | --- | --- | --- |
| 1 | exp eye disease/ | 1096631 | Advanced |
| 2 | exp hereditary optic atrophy/ | 5029 | Advanced |
| 3 | visual field/ | 38202 | Advanced |
| 4 | exp vision test/ | 43705 | Advanced |
| 5 | exp hearing/ | 68535 | Advanced |
| 6 | sound detection/ | 11589 | Advanced |
| 7 | (eye adj2 disease*).mp. | 65024 | Advanced |
| 8 | (eye adj2 disorder*).mp. | 8536 | Advanced |
| 9 | Albinism.mp. | 6862 | Advanced |
| 10 | (Chediak adj2 higashi syndrome).mp. | 1745 | Advanced |
| 11 | (Congenital adj2 amauros?s).mp. | 2804 | Advanced |
| 12 | dysgenesis neuroepithelialis retinae.mp. | 0 | Advanced |
| 13 | hereditary epithelial dysplasia of retina.mp. | 0 | Advanced |
| 14 | hereditary retinal aplasia.mp. | 0 | Advanced |
| 15 | heredoretinopathia congenitalis.mp. | 0 | Advanced |
| 16 | leber abiotroph*.mp. | 0 | Advanced |
| 17 | (leber* adj2 amauros?s).mp. | 2812 | Advanced |
| 18 | leber congenital tapetoretinal degeneration.mp. | 0 | Advanced |
| 19 | hereditary optic atroph*.mp. | 1052 | Advanced |
| 20 | (optic adj2 hypoplasia*).mp. | 1675 | Advanced |
| 21 | (retinal adj2 degeneration*).mp. | 14352 | Advanced |
| 22 | (retinal adj2 dysplasia*).mp. | 433 | Advanced |
| 23 | pigmentary retinopathy*.mp. | 830 | Advanced |
| 24 | retinitis pigmentosa.mp. | 16282 | Advanced |
| 25 | tapetoretinal degeneration*.mp. | 608 | Advanced |
| 26 | fundus flavimaculatus.mp. | 274 | Advanced |
| 27 | macular dystrophy with flecks.mp. | 2 | Advanced |
| 28 | stargardt disease.mp. | 2380 | Advanced |
| 29 | cod md syndrome*.mp. | 3 | Advanced |
| 30 | cerebromuscular dystrophy.mp. | 14 | Advanced |
| 31 | cerebroocular dysplasia muscular dystrophy syndrome.mp. | 0 | Advanced |
| 32 | chemke syndrome.mp. | 1 | Advanced |
| 33 | congenital muscular dystrophy dystroglycanopathy.mp. | 13 | Advanced |
| 34 | walker-warburg syndrome*.mp. | 876 | Advanced |
| 35 | fukuyama cmd.mp. | 42 | Advanced |
| 36 | (fukuyama adj3 muscular dystrophy).mp. | 700 | Advanced |
| 37 | fukuyama syndrome.mp. | 2 | Advanced |
| 38 | hard syndrome*.mp. | 5 | Advanced |
| 39 | lgmd2k.mp. | 10 | Advanced |
| 40 | mddga1.mp. | 2 | Advanced |
| 41 | meb syndrome.mp. | 0 | Advanced |
| 42 | muscle-eye-brain syndrome.mp. | 3 | Advanced |
| 43 | muscle eye brain disease*.mp. | 399 | Advanced |
| 44 | muscular dystrophy due to defective glycosylation of dystroglycan 4a.mp. | 0 | Advanced |
| 45 | muscular dystrophy-dystroglycanopathy.mp. | 55 | Advanced |
| 46 | pagon syndrome*.mp. | 0 | Advanced |
| 47 | warburg syndrome.mp. | 885 | Advanced |
| 48 | alpha dystroglycanopathies.mp. | 91 | Advanced |
| 49 | congenital mesodermal dysmorphodystroph*.mp. | 0 | Advanced |
| 50 | gems.mp. | 1866 | Advanced |
| 51 | glaucoma-lens ectopia-microspherophakia-stiffness-shortness syndrome.mp. | 1 | Advanced |
| 52 | (marchesani adj2 syndrome*).mp. | 312 | Advanced |
| 53 | spherophakia brachymorphia syndrome*.mp. | 2 | Advanced |
| 54 | cataract*.mp. | 109335 | Advanced |
| 55 | lens opacity*.mp. | 1389 | Advanced |
| 56 | pseudoaphakia*.mp. | 10 | Advanced |
| 57 | (lens adj2 cloud*).mp. | 84 | Advanced |
| 58 | glaucoma*.mp. | 109013 | Advanced |
| 59 | increased intraocular pressure.mp. | 1734 | Advanced |
| 60 | ocular hypertension.mp. | 7846 | Advanced |
| 61 | optic neuropathy.mp. | 18337 | Advanced |
| 62 | brown* tendon sheath syndrome*.mp. | 3 | Advanced |
| 63 | conjugate gaze spasm*.mp. | 0 | Advanced |
| 64 | convergence excess*.mp. | 121 | Advanced |
| 65 | convergence insufficienc*.mp. | 680 | Advanced |
| 66 | cyclophoria*.mp. | 23 | Advanced |
| 67 | eye motility disorder*.mp. | 24 | Advanced |
| 68 | eye movement disorder*.mp. | 6176 | Advanced |
| 69 | internuclear ophthalmoplegia*.mp. | 1386 | Advanced |
| 70 | ocular motility disorder*.mp. | 512 | Advanced |
| 71 | ocular torticollis.mp. | 94 | Advanced |
| 72 | opsoclonus.mp. | 2032 | Advanced |
| 73 | parinaud* syndrome*.mp. | 469 | Advanced |
| 74 | paroxysmal ocular dyskinesia*.mp. | 0 | Advanced |
| 75 | pseudoophthalmoplegia*.mp. | 1 | Advanced |
| 76 | skew deviation*.mp. | 507 | Advanced |
| 77 | smooth pursuit deficienc*.mp. | 2 | Advanced |
| 78 | spasm of conjugate gaze.mp. | 1 | Advanced |
| 79 | tendon sheath syndrome of brown.mp. | 4 | Advanced |
| 80 | fisher syndrome.mp. | 1773 | Advanced |
| 81 | miller fisher.mp. | 1638 | Advanced |
| 82 | (ophthalmoplegia, ataxia and areflexia syndrome).mp. | 5 | Advanced |
| 83 | nystagmus.mp. | 30521 | Advanced |
| 84 | involuntary eye movement*.mp. | 193 | Advanced |
| 85 | Jerky eye movement*.mp. | 28 | Advanced |
| 86 | cranial nerve iii disease*.mp. | 0 | Advanced |
| 87 | oculomotor nerve disease*.mp. | 1313 | Advanced |
| 88 | oculomotor nerve disorder*.mp. | 3 | Advanced |
| 89 | oculomotor nerve pals*.mp. | 1291 | Advanced |
| 90 | oculomotor nerve paralys?s.mp. | 95 | Advanced |
| 91 | oculomotor neuropath*.mp. | 28 | Advanced |
| 92 | third cranial nerve disease*.mp. | 0 | Advanced |
| 93 | third nerve pals*.mp. | 1072 | Advanced |
| 94 | third nerve paralysis.mp. | 39 | Advanced |
| 95 | ophthalmoplegia*.mp. | 18915 | Advanced |
| 96 | oculomotor paralysis.mp. | 210 | Advanced |
| 97 | ophthalmopares?s.mp. | 905 | Advanced |
| 98 | dancing eyes.mp. | 60 | Advanced |
| 99 | myoclonic encephalopathy*.mp. | 410 | Advanced |
| 100 | kinsbourne syndrome.mp. | 58 | Advanced |
| 101 | opsoclonus myoclonus.mp. | 1375 | Advanced |
| 102 | strabismus.mp. | 31851 | Advanced |
| 103 | dissociated horizontal deviation*.mp. | 36 | Advanced |
| 104 | dissociated vertical deviation*.mp. | 373 | Advanced |
| 105 | heterophoria*.mp. | 520 | Advanced |
| 106 | heterotropia*.mp. | 151 | Advanced |
| 107 | hypertropia*.mp. | 639 | Advanced |
| 108 | phoria*.mp. | 659 | Advanced |
| 109 | squint*.mp. | 2158 | Advanced |
| 110 | crossed eye*.mp. | 40 | Advanced |
| 111 | tolosa hunt syndrome*.mp. | 983 | Advanced |
| 112 | Ametropia*.mp. | 1484 | Advanced |
| 113 | refractive disorder*.mp. | 107 | Advanced |
| 114 | refractive error*.mp. | 14732 | Advanced |
| 115 | aniseikonia.mp. | 556 | Advanced |
| 116 | anisometropia.mp. | 3917 | Advanced |
| 117 | astigmatism*.mp. | 19393 | Advanced |
| 118 | corneal wavefront aberration*.mp. | 677 | Advanced |
| 119 | farsighted*.mp. | 170 | Advanced |
| 120 | longsighted*.mp. | 9 | Advanced |
| 121 | hypermetropia*.mp. | 7952 | Advanced |
| 122 | hyperopia*.mp. | 4394 | Advanced |
| 123 | myopia*.mp. | 37590 | Advanced |
| 124 | nearsighted*.mp. | 185 | Advanced |
| 125 | shortsighted*.mp. | 166 | Advanced |
| 126 | presbyopia*.mp. | 3471 | Advanced |
| 127 | (retinal adj2 disease*).mp. | 15371 | Advanced |
| 128 | (cone adj2 dystroph*).mp. | 2594 | Advanced |
| 129 | diabetic retinopath*.mp. | 61957 | Advanced |
| 130 | hypertensive retinopath*.mp. | 1164 | Advanced |
| 131 | diabetic eye disease*.mp. | 1012 | Advanced |
| 132 | diabetic macular edema.mp. | 10166 | Advanced |
| 133 | blindness.mp. | 72791 | Advanced |
| 134 | hemeralopia*.mp. | 88 | Advanced |
| 135 | macropsia*.mp. | 122 | Advanced |
| 136 | metamorphopsia*.mp. | 1685 | Advanced |
| 137 | micropsia*.mp. | 200 | Advanced |
| 138 | vision disabilit*.mp. | 97 | Advanced |
| 139 | vision disorder*.mp. | 1587 | Advanced |
| 140 | vision impairment*.mp. | 3878 | Advanced |
| 141 | visual disorder*.mp. | 34387 | Advanced |
| 142 | visual impairment*.mp. | 71592 | Advanced |
| 143 | visual disabilit*.mp. | 1392 | Advanced |
| 144 | amblyopia*.mp. | 13511 | Advanced |
| 145 | lazy eye*.mp. | 95 | Advanced |
| 146 | amauros?s.mp. | 5204 | Advanced |
| 147 | sudden visual loss*.mp. | 381 | Advanced |
| 148 | achromatopsia*.mp. | 920 | Advanced |
| 149 | colo?r vision defect*.mp. | 4063 | Advanced |
| 150 | colo?r vision deficienc*.mp. | 702 | Advanced |
| 151 | colo?r vision impairment*.mp. | 133 | Advanced |
| 152 | colo?r perception deficienc*.mp. | 3 | Advanced |
| 153 | deutan defect.mp. | 10 | Advanced |
| 154 | monochromatopsia.mp. | 1 | Advanced |
| 155 | protan defect.mp. | 15 | Advanced |
| 156 | tritan defect.mp. | 33 | Advanced |
| 157 | diplopia*.mp. | 31757 | Advanced |
| 158 | double vision.mp. | 2262 | Advanced |
| 159 | polyopsia*.mp. | 3 | Advanced |
| 160 | nyctalopia.mp. | 571 | Advanced |
| 161 | light sensitivit*.mp. | 2640 | Advanced |
| 162 | photophobia*.mp. | 15061 | Advanced |
| 163 | scotoma*.mp. | 8691 | Advanced |
| 164 | retinocochleocerebral vasculopath*.mp. | 21 | Advanced |
| 165 | susac* syndrome.mp. | 834 | Advanced |
| 166 | diminished vision.mp. | 206 | Advanced |
| 167 | reduced vision.mp. | 1112 | Advanced |
| 168 | subnormal vision.mp. | 88 | Advanced |
| 169 | sub-normal vision.mp. | 7 | Advanced |
| 170 | low vision.mp. | 8283 | Advanced |
| 171 | macular degeneration*.mp. | 41832 | Advanced |
| 172 | maculopath*.mp. | 11635 | Advanced |
| 173 | macular dystroph*.mp. | 2309 | Advanced |
| 174 | macular disorder*.mp. | 228 | Advanced |
| 175 | retinal degeneration*.mp. | 12875 | Advanced |
| 176 | retinal degenerative disease*.mp. | 1547 | Advanced |
| 177 | visual field*.mp. | 63957 | Advanced |
| 178 | vision test*.mp. | 12352 | Advanced |
| 179 | colo?r perception test*.mp. | 41 | Advanced |
| 180 | ocular refraction*.mp. | 263 | Advanced |
| 181 | vision screening.mp. | 1799 | Advanced |
| 182 | visual acuit*.mp. | 172087 | Advanced |
| 183 | visual contrast sensitivity*.mp. | 295 | Advanced |
| 184 | emmetropia*.mp. | 2944 | Advanced |
| 185 | automated perimetry exam*.mp. | 9 | Advanced |
| 186 | campimetr*.mp. | 379 | Advanced |
| 187 | perimetr*.mp. | 18089 | Advanced |
| 188 | tangent screen exam*.mp. | 12 | Advanced |
| 189 | auditory perception.mp. | 3073 | Advanced |
| 190 | auditory processing.mp. | 6601 | Advanced |
| 191 | (auditory adj2 localization*).mp. | 525 | Advanced |
| 192 | (sound adj2 localization*).mp. | 2769 | Advanced |
| 193 | spatial hearing.mp. | 620 | Advanced |
| 194 | or/1-193 | 1291841 | Advanced |
| 195 | virtual reality/ | 26318 | Advanced |
| 196 | augmented reality/ | 2278 | Advanced |
| 197 | (virtual adj3 realit*).mp. | 33269 | Advanced |
| 198 | (augmented adj3 realit*).mp. | 5671 | Advanced |
| 199 | VRET.mp. | 169 | Advanced |
| 200 | iVR.mp. | 2748 | Advanced |
| 201 | iVRs.mp. | 390 | Advanced |
| 202 | iVE.mp. | 3225 | Advanced |
| 203 | iVEs.mp. | 413 | Advanced |
| 204 | (immers* adj2 environment*).mp. | 905 | Advanced |
| 205 | samsung gear.mp. | 72 | Advanced |
| 206 | oculus rift.mp. | 273 | Advanced |
| 207 | HCD vive.mp. | 0 | Advanced |
| 208 | hololens.mp. | 543 | Advanced |
| 209 | gear VR.mp. | 57 | Advanced |
| 210 | head-mount*.mp. | 3867 | Advanced |
| 211 | (head adj3 gear*).mp. | 124 | Advanced |
| 212 | HMD.mp. | 1733 | Advanced |
| 213 | Gen2-VR.mp. | 1 | Advanced |
| 214 | (google adj2 daydream).mp. | 8 | Advanced |
| 215 | (google adj2 cardboard).mp. | 43 | Advanced |
| 216 | PlayStation.mp. | 141 | Advanced |
| 217 | (ReTrak adj2 Utopia).mp. | 0 | Advanced |
| 218 | NOON VR.mp. | 0 | Advanced |
| 219 | Freefly VR.mp. | 0 | Advanced |
| 220 | Homido.mp. | 0 | Advanced |
| 221 | iWear.mp. | 5 | Advanced |
| 222 | (mixed adj2 realit*).mp. | 1074 | Advanced |
| 223 | (simulat* adj2 environment*).mp. | 6869 | Advanced |
| 224 | (virtual adj2 environment*).mp. | 8432 | Advanced |
| 225 | exp video game/ | 6390 | Advanced |
| 226 | (computer adj2 game*).mp. | 2430 | Advanced |
| 227 | (video adj2 game*).mp. | 9519 | Advanced |
| 228 | gaming.mp. | 6856 | Advanced |
| 229 | computer interface/ | 36207 | Advanced |
| 230 | (user adj2 interface*).mp. | 12794 | Advanced |
| 231 | (virtual adj2 interface*).mp. | 234 | Advanced |
| 232 | Oculus Quest.mp. | 70 | Advanced |
| 233 | Meta Quest.mp. | 14 | Advanced |
| 234 | Metaverse.mp. | 258 | Advanced |
| 235 | (simulate* adj2 realit*).mp. | 141 | Advanced |
| 236 | (tethered adj2 headset*).mp. | 2 | Advanced |
| 237 | Serious game.mp. | 758 | Advanced |
| 238 | or/195-237 | 110501 | Advanced |
| 239 | 194 and 238 | 4422 | Advanced |
| 240 | (exp animals/ or exp animal experimentation/ or nonhuman/) not ((exp animals/ or exp animal experimentation/ or nonhuman/) and exp human/) | 7233074 | Advanced |
| 241 | 239 not 240 | 4267 | Advanced |
| 242 | limit 241 to english language | 4121 | Advanced |
| 243 | limit 242 to (conference abstract or conference paper or "conference review" or conference proceeding) | 827 | Advanced |
| 244 | 242 not 243 | 3294 | Advanced |
| 245 | remove duplicates from 244 | 3221 | Advanced |

EBM Reviews - Cochrane Central Register of Controlled Trials

| **#** | **Searches** | **Results** | **Type** |
| --- | --- | --- | --- |
| 1 | exp eye diseases/ | 25051 | Advanced |
| 2 | Optic Atrophy/ge | 0 | Advanced |
| 3 | exp Optic Nerve/ab | 0 | Advanced |
| 4 | Visual Fields/ | 1023 | Advanced |
| 5 | exp Vision Tests/ | 7632 | Advanced |
| 6 | exp Auditory Perception/ | 2280 | Advanced |
| 7 | Sound Localization/ | 104 | Advanced |
| 8 | (eye adj2 disease*).mp. | 4453 | Advanced |
| 9 | (eye adj2 disorder*).mp. | 526 | Advanced |
| 10 | Albinism.mp. | 24 | Advanced |
| 11 | (Chediak adj2 higashi syndrome).mp. | 2 | Advanced |
| 12 | (Congenital adj2 amauros?s).mp. | 31 | Advanced |
| 13 | dysgenesis neuroepithelialis retinae.mp. | 0 | Advanced |
| 14 | hereditary epithelial dysplasia of retina.mp. | 0 | Advanced |
| 15 | hereditary retinal aplasia.mp. | 0 | Advanced |
| 16 | heredoretinopathia congenitalis.mp. | 0 | Advanced |
| 17 | leber abiotroph*.mp. | 0 | Advanced |
| 18 | (leber* adj2 amauros?s).mp. | 30 | Advanced |
| 19 | leber congenital tapetoretinal degeneration.mp. | 0 | Advanced |
| 20 | hereditary optic atroph*.mp. | 2 | Advanced |
| 21 | (optic adj2 hypoplasia*).mp. | 9 | Advanced |
| 22 | (retinal adj2 degeneration*).mp. | 145 | Advanced |
| 23 | (retinal adj2 dysplasia*).mp. | 0 | Advanced |
| 24 | pigmentary retinopathy*.mp. | 3 | Advanced |
| 25 | retinitis pigmentosa.mp. | 307 | Advanced |
| 26 | tapetoretinal degeneration*.mp. | 0 | Advanced |
| 27 | fundus flavimaculatus.mp. | 5 | Advanced |
| 28 | macular dystrophy with flecks.mp. | 0 | Advanced |
| 29 | stargardt disease.mp. | 54 | Advanced |
| 30 | cod md syndrome*.mp. | 0 | Advanced |
| 31 | cerebromuscular dystrophy.mp. | 0 | Advanced |
| 32 | cerebroocular dysplasia muscular dystrophy syndrome.mp. | 0 | Advanced |
| 33 | chemke syndrome.mp. | 0 | Advanced |
| 34 | congenital muscular dystrophy dystroglycanopathy.mp. | 0 | Advanced |
| 35 | walker-warburg syndrome*.mp. | 0 | Advanced |
| 36 | fukuyama cmd.mp. | 0 | Advanced |
| 37 | (fukuyama adj3 muscular dystrophy).mp. | 7 | Advanced |
| 38 | fukuyama syndrome.mp. | 0 | Advanced |
| 39 | hard syndrome*.mp. | 0 | Advanced |
| 40 | lgmd2k.mp. | 0 | Advanced |
| 41 | mddga1.mp. | 0 | Advanced |
| 42 | meb syndrome.mp. | 0 | Advanced |
| 43 | muscle-eye-brain syndrome.mp. | 0 | Advanced |
| 44 | muscle eye brain disease*.mp. | 1 | Advanced |
| 45 | muscular dystrophy due to defective glycosylation of dystroglycan 4a.mp. | 0 | Advanced |
| 46 | muscular dystrophy-dystroglycanopathy.mp. | 0 | Advanced |
| 47 | pagon syndrome*.mp. | 0 | Advanced |
| 48 | warburg syndrome.mp. | 0 | Advanced |
| 49 | alpha dystroglycanopathies.mp. | 0 | Advanced |
| 50 | congenital mesodermal dysmorphodystroph*.mp. | 0 | Advanced |
| 51 | gems.mp. | 93 | Advanced |
| 52 | glaucoma-lens ectopia-microspherophakia-stiffness-shortness syndrome.mp. | 0 | Advanced |
| 53 | (marchesani adj2 syndrome*).mp. | 0 | Advanced |
| 54 | spherophakia brachymorphia syndrome*.mp. | 0 | Advanced |
| 55 | cataract*.mp. | 8936 | Advanced |
| 56 | lens opacity*.mp. | 121 | Advanced |
| 57 | pseudoaphakia*.mp. | 0 | Advanced |
| 58 | (lens adj2 cloud*).mp. | 10 | Advanced |
| 59 | glaucoma*.mp. | 8786 | Advanced |
| 60 | increased intraocular pressure.mp. | 158 | Advanced |
| 61 | ocular hypertension.mp. | 2676 | Advanced |
| 62 | optic neuropathy.mp. | 507 | Advanced |
| 63 | brown* tendon sheath syndrome*.mp. | 0 | Advanced |
| 64 | conjugate gaze spasm*.mp. | 0 | Advanced |
| 65 | convergence excess*.mp. | 8 | Advanced |
| 66 | convergence insufficienc*.mp. | 120 | Advanced |
| 67 | cyclophoria*.mp. | 1 | Advanced |
| 68 | eye motility disorder*.mp. | 0 | Advanced |
| 69 | eye movement disorder*.mp. | 50 | Advanced |
| 70 | internuclear ophthalmoplegia*.mp. | 7 | Advanced |
| 71 | ocular motility disorder*.mp. | 110 | Advanced |
| 72 | ocular torticollis.mp. | 1 | Advanced |
| 73 | opsoclonus.mp. | 14 | Advanced |
| 74 | parinaud* syndrome*.mp. | 1 | Advanced |
| 75 | paroxysmal ocular dyskinesia*.mp. | 0 | Advanced |
| 76 | pseudoophthalmoplegia*.mp. | 0 | Advanced |
| 77 | skew deviation*.mp. | 4 | Advanced |
| 78 | smooth pursuit deficienc*.mp. | 0 | Advanced |
| 79 | spasm of conjugate gaze.mp. | 0 | Advanced |
| 80 | tendon sheath syndrome of brown.mp. | 0 | Advanced |
| 81 | fisher syndrome.mp. | 5 | Advanced |
| 82 | miller fisher.mp. | 3 | Advanced |
| 83 | (ophthalmoplegia, ataxia and areflexia syndrome).mp. [mp=title, original title, abstract, floating sub-heading word, mesh headings, heading words, keyword] | 0 | Advanced |
| 84 | nystagmus.mp. | 660 | Advanced |
| 85 | involuntary eye movement*.mp. | 7 | Advanced |
| 86 | Jerky eye movement*.mp. | 0 | Advanced |
| 87 | cranial nerve iii disease*.mp. | 0 | Advanced |
| 88 | oculomotor nerve disease*.mp. | 11 | Advanced |
| 89 | oculomotor nerve disorder*.mp. | 0 | Advanced |
| 90 | oculomotor nerve pals*.mp. | 9 | Advanced |
| 91 | oculomotor nerve paralys?s.mp. | 0 | Advanced |
| 92 | oculomotor neuropath*.mp. | 0 | Advanced |
| 93 | third cranial nerve disease*.mp. | 0 | Advanced |
| 94 | third nerve pals*.mp. | 2 | Advanced |
| 95 | third nerve paralysis.mp. | 0 | Advanced |
| 96 | ophthalmoplegia*.mp. | 87 | Advanced |
| 97 | oculomotor paralysis.mp. | 5 | Advanced |
| 98 | ophthalmopares?s.mp. | 6 | Advanced |
| 99 | dancing eyes.mp. | 0 | Advanced |
| 100 | myoclonic encephalopathy*.mp. | 2 | Advanced |
| 101 | kinsbourne syndrome.mp. | 0 | Advanced |
| 102 | opsoclonus myoclonus.mp. | 13 | Advanced |
| 103 | strabismus.mp. | 1370 | Advanced |
| 104 | dissociated horizontal deviation*.mp. | 0 | Advanced |
| 105 | dissociated vertical deviation*.mp. | 16 | Advanced |
| 106 | heterophoria*.mp. | 61 | Advanced |
| 107 | heterotropia*.mp. | 14 | Advanced |
| 108 | hypertropia*.mp. | 15 | Advanced |
| 109 | phoria*.mp. | 123 | Advanced |
| 110 | squint*.mp. | 143 | Advanced |
| 111 | crossed eye*.mp. | 1 | Advanced |
| 112 | tolosa hunt syndrome*.mp. | 1 | Advanced |
| 113 | Ametropia*.mp. | 126 | Advanced |
| 114 | refractive disorder*.mp. | 4 | Advanced |
| 115 | refractive error*.mp. | 1789 | Advanced |
| 116 | aniseikonia.mp. | 21 | Advanced |
| 117 | anisometropia.mp. | 334 | Advanced |
| 118 | astigmatism*.mp. | 2327 | Advanced |
| 119 | corneal wavefront aberration*.mp. | 141 | Advanced |
| 120 | farsighted*.mp. | 11 | Advanced |
| 121 | longsighted*.mp. | 0 | Advanced |
| 122 | hypermetropia*.mp. | 144 | Advanced |
| 123 | hyperopia*.mp. | 308 | Advanced |
| 124 | myopia*.mp. | 3336 | Advanced |
| 125 | nearsighted*.mp. | 47 | Advanced |
| 126 | shortsighted*.mp. | 11 | Advanced |
| 127 | presbyopia*.mp. | 396 | Advanced |
| 128 | (retinal adj2 disease*).mp. | 957 | Advanced |
| 129 | (cone adj2 dystroph*).mp. | 24 | Advanced |
| 130 | diabetic retinopath*.mp. | 4442 | Advanced |
| 131 | hypertensive retinopath*.mp. | 27 | Advanced |
| 132 | diabetic eye disease*.mp. | 92 | Advanced |
| 133 | diabetic macular edema.mp. | 1783 | Advanced |
| 134 | blindness.mp. | 2608 | Advanced |
| 135 | hemeralopia*.mp. | 0 | Advanced |
| 136 | macropsia*.mp. | 0 | Advanced |
| 137 | metamorphopsia*.mp. | 99 | Advanced |
| 138 | micropsia*.mp. | 3 | Advanced |
| 139 | vision disabilit*.mp. | 2 | Advanced |
| 140 | vision disorder*.mp. | 908 | Advanced |
| 141 | vision impairment*.mp. | 255 | Advanced |
| 142 | visual disorder*.mp. | 667 | Advanced |
| 143 | visual impairment*.mp. | 2034 | Advanced |
| 144 | visual disabilit*.mp. | 66 | Advanced |
| 145 | amblyopia*.mp. | 822 | Advanced |
| 146 | lazy eye*.mp. | 26 | Advanced |
| 147 | amauros?s.mp. | 90 | Advanced |
| 148 | sudden visual loss*.mp. | 2 | Advanced |
| 149 | achromatopsia*.mp. | 14 | Advanced |
| 150 | colo?r vision defect*.mp. | 90 | Advanced |
| 151 | colo?r vision deficienc*.mp. | 22 | Advanced |
| 152 | colo?r vision impairment*.mp. | 7 | Advanced |
| 153 | colo?r perception deficienc*.mp. | 0 | Advanced |
| 154 | deutan defect.mp. | 1 | Advanced |
| 155 | monochromatopsia.mp. | 0 | Advanced |
| 156 | protan defect.mp. | 2 | Advanced |
| 157 | tritan defect.mp. | 1 | Advanced |
| 158 | diplopia*.mp. | 687 | Advanced |
| 159 | double vision.mp. | 90 | Advanced |
| 160 | polyopsia*.mp. | 0 | Advanced |
| 161 | nyctalopia.mp. | 4 | Advanced |
| 162 | light sensitivit*.mp. | 161 | Advanced |
| 163 | photophobia*.mp. | 1201 | Advanced |
| 164 | scotoma*.mp. | 273 | Advanced |
| 165 | retinocochleocerebral vasculopath*.mp. | 0 | Advanced |
| 166 | susac* syndrome.mp. | 1 | Advanced |
| 167 | diminished vision.mp. | 13 | Advanced |
| 168 | reduced vision.mp. | 73 | Advanced |
| 169 | subnormal vision.mp. | 3 | Advanced |
| 170 | sub-normal vision.mp. | 0 | Advanced |
| 171 | low vision.mp. | 505 | Advanced |
| 172 | macular degeneration*.mp. | 3756 | Advanced |
| 173 | maculopath*.mp. | 436 | Advanced |
| 174 | macular dystroph*.mp. | 33 | Advanced |
| 175 | macular disorder*.mp. | 9 | Advanced |
| 176 | retinal degeneration*.mp. | 106 | Advanced |
| 177 | retinal degenerative disease*.mp. | 17 | Advanced |
| 178 | visual field*.mp. | 3679 | Advanced |
| 179 | vision test*.mp. | 658 | Advanced |
| 180 | colo?r perception test*.mp. | 52 | Advanced |
| 181 | ocular refraction*.mp. | 12 | Advanced |
| 182 | vision screening.mp. | 238 | Advanced |
| 183 | visual acuit*.mp. | 16976 | Advanced |
| 184 | visual contrast sensitivity*.mp. | 29 | Advanced |
| 185 | emmetropia*.mp. | 167 | Advanced |
| 186 | automated perimetry exam*.mp. | 0 | Advanced |
| 187 | campimetr*.mp. | 17 | Advanced |
| 188 | perimetr*.mp. | 1101 | Advanced |
| 189 | tangent screen exam*.mp. | 1 | Advanced |
| 190 | auditory perception.mp. | 917 | Advanced |
| 191 | auditory processing.mp. | 284 | Advanced |
| 192 | (auditory adj2 localization*).mp. | 14 | Advanced |
| 193 | (sound adj2 localization*).mp. | 132 | Advanced |
| 194 | spatial hearing.mp. | 28 | Advanced |
| 195 | or/1-194 | 55231 | Advanced |
| 196 | Virtual Reality/ | 982 | Advanced |
| 197 | (virtual adj3 realit*).mp. | 6017 | Advanced |
| 198 | (augmented adj3 realit*).mp. | 472 | Advanced |
| 199 | VRET.mp. | 98 | Advanced |
| 200 | iVR.mp. | 668 | Advanced |
| 201 | iVRs.mp. | 189 | Advanced |
| 202 | iVE.mp. | 148 | Advanced |
| 203 | iVEs.mp. | 27 | Advanced |
| 204 | (immers* adj2 environment*).mp. | 138 | Advanced |
| 205 | samsung gear.mp. | 15 | Advanced |
| 206 | oculus rift.mp. | 62 | Advanced |
| 207 | HCD vive.mp. | 0 | Advanced |
| 208 | hololens.mp. | 46 | Advanced |
| 209 | gear VR.mp. | 11 | Advanced |
| 210 | head-mount*.mp. | 469 | Advanced |
| 211 | (head adj3 gear*).mp. | 19 | Advanced |
| 212 | HMD.mp. | 215 | Advanced |
| 213 | Gen2-VR.mp. | 0 | Advanced |
| 214 | (google adj2 daydream).mp. | 2 | Advanced |
| 215 | (google adj2 cardboard).mp. | 13 | Advanced |
| 216 | PlayStation.mp. | 32 | Advanced |
| 217 | (ReTrak adj2 Utopia).mp. | 0 | Advanced |
| 218 | NOON VR.mp. | 1 | Advanced |
| 219 | Freefly VR.mp. | 0 | Advanced |
| 220 | Homido.mp. | 0 | Advanced |
| 221 | iWear.mp. | 0 | Advanced |
| 222 | (mixed adj2 realit*).mp. | 121 | Advanced |
| 223 | (simulat* adj2 environment*).mp. | 422 | Advanced |
| 224 | (virtual adj2 environment*).mp. | 911 | Advanced |
| 225 | Video Games/ | 1000 | Advanced |
| 226 | (computer adj2 game*).mp. | 665 | Advanced |
| 227 | (video adj2 game*).mp. | 2356 | Advanced |
| 228 | gaming.mp. | 979 | Advanced |
| 229 | user-computer interface/ | 1422 | Advanced |
| 230 | (user adj2 interface*).mp. | 1742 | Advanced |
| 231 | (virtual adj2 interface*).mp. | 21 | Advanced |
| 232 | Oculus Quest.mp. | 36 | Advanced |
| 233 | Meta Quest.mp. | 7 | Advanced |
| 234 | Metaverse.mp. | 10 | Advanced |
| 235 | (simulate* adj2 realit*).mp. | 14 | Advanced |
| 236 | (tethered adj2 headset*).mp. | 0 | Advanced |
| 237 | Serious game.mp. | 266 | Advanced |
| 238 | or/196-237 | 12222 | Advanced |
| 239 | 195 and 238 | 569 | Advanced |
| 240 | remove duplicates from 239 | 566 | Advanced |

EBM Reviews - Cochrane Database of Systematic Reviews <2005 to August 9, 2023>

| **#** | **Searches** | **Results** | **Type** |
| --- | --- | --- | --- |
| 1 | (eye adj2 disease*).ti,ab. | 16 | Advanced |
| 2 | (eye adj2 disorder*).ti,ab. | 3 | Advanced |
| 3 | Albinism.ti,ab. | 0 | Advanced |
| 4 | (Chediak adj2 higashi syndrome).ti,ab. | 0 | Advanced |
| 5 | (Congenital adj2 amauros?s).ti,ab. | 0 | Advanced |
| 6 | dysgenesis neuroepithelialis retinae.ti,ab. | 0 | Advanced |
| 7 | hereditary epithelial dysplasia of retina.ti,ab. | 0 | Advanced |
| 8 | hereditary retinal aplasia.ti,ab. | 0 | Advanced |
| 9 | heredoretinopathia congenitalis.ti,ab. | 0 | Advanced |
| 10 | leber abiotroph*.ti,ab. | 0 | Advanced |
| 11 | (leber* adj2 amauros?s).ti,ab. | 0 | Advanced |
| 12 | leber congenital tapetoretinal degeneration.ti,ab. | 0 | Advanced |
| 13 | hereditary optic atroph*.ti,ab. | 0 | Advanced |
| 14 | (optic adj2 hypoplasia*).ti,ab. | 0 | Advanced |
| 15 | (retinal adj2 degeneration*).ti,ab. | 1 | Advanced |
| 16 | (retinal adj2 dysplasia*).ti,ab. | 0 | Advanced |
| 17 | pigmentary retinopathy*.ti,ab. | 0 | Advanced |
| 18 | retinitis pigmentosa.ti,ab. | 1 | Advanced |
| 19 | tapetoretinal degeneration*.ti,ab. | 0 | Advanced |
| 20 | fundus flavimaculatus.ti,ab. | 0 | Advanced |
| 21 | macular dystrophy with flecks.ti,ab. | 0 | Advanced |
| 22 | stargardt disease.ti,ab. | 0 | Advanced |
| 23 | cod md syndrome*.ti,ab. | 0 | Advanced |
| 24 | cerebromuscular dystrophy.ti,ab. | 0 | Advanced |
| 25 | cerebroocular dysplasia muscular dystrophy syndrome.ti,ab. | 0 | Advanced |
| 26 | chemke syndrome.ti,ab. | 0 | Advanced |
| 27 | congenital muscular dystrophy dystroglycanopathy.ti,ab. | 0 | Advanced |
| 28 | walker-warburg syndrome*.ti,ab. | 0 | Advanced |
| 29 | fukuyama cmd.ti,ab. | 0 | Advanced |
| 30 | (fukuyama adj3 muscular dystrophy).ti,ab. | 0 | Advanced |
| 31 | fukuyama syndrome.ti,ab. | 0 | Advanced |
| 32 | hard syndrome*.ti,ab. | 0 | Advanced |
| 33 | lgmd2k.ti,ab. | 0 | Advanced |
| 34 | mddga1.ti,ab. | 0 | Advanced |
| 35 | meb syndrome.ti,ab. | 0 | Advanced |
| 36 | muscle-eye-brain syndrome.ti,ab. | 0 | Advanced |
| 37 | muscle eye brain disease*.ti,ab. | 0 | Advanced |
| 38 | muscular dystrophy due to defective glycosylation of dystroglycan 4a.ti,ab. | 0 | Advanced |
| 39 | muscular dystrophy-dystroglycanopathy.ti,ab. | 0 | Advanced |
| 40 | pagon syndrome*.ti,ab. | 0 | Advanced |
| 41 | warburg syndrome.ti,ab. | 0 | Advanced |
| 42 | alpha dystroglycanopathies.ti,ab. | 0 | Advanced |
| 43 | congenital mesodermal dysmorphodystroph*.ti,ab. | 0 | Advanced |
| 44 | gems.ti,ab. | 1 | Advanced |
| 45 | glaucoma-lens ectopia-microspherophakia-stiffness-shortness syndrome.ti,ab. | 0 | Advanced |
| 46 | (marchesani adj2 syndrome*).ti,ab. | 0 | Advanced |
| 47 | spherophakia brachymorphia syndrome*.ti,ab. | 0 | Advanced |
| 48 | cataract*.ti,ab. | 73 | Advanced |
| 49 | lens opacity*.ti,ab. | 2 | Advanced |
| 50 | pseudoaphakia*.ti,ab. | 0 | Advanced |
| 51 | (lens adj2 cloud*).ti,ab. | 0 | Advanced |
| 52 | glaucoma*.ti,ab. | 61 | Advanced |
| 53 | increased intraocular pressure.ti,ab. | 4 | Advanced |
| 54 | ocular hypertension.ti,ab. | 11 | Advanced |
| 55 | optic neuropathy.ti,ab. | 15 | Advanced |
| 56 | brown* tendon sheath syndrome*.ti,ab. | 0 | Advanced |
| 57 | conjugate gaze spasm*.ti,ab. | 0 | Advanced |
| 58 | convergence excess*.ti,ab. | 0 | Advanced |
| 59 | convergence insufficienc*.ti,ab. | 1 | Advanced |
| 60 | cyclophoria*.ti,ab. | 0 | Advanced |
| 61 | eye motility disorder*.ti,ab. | 0 | Advanced |
| 62 | eye movement disorder*.ti,ab. | 3 | Advanced |
| 63 | internuclear ophthalmoplegia*.ti,ab. | 0 | Advanced |
| 64 | ocular motility disorder*.ti,ab. | 0 | Advanced |
| 65 | ocular torticollis.ti,ab. | 1 | Advanced |
| 66 | opsoclonus.ti,ab. | 0 | Advanced |
| 67 | parinaud* syndrome*.ti,ab. | 0 | Advanced |
| 68 | paroxysmal ocular dyskinesia*.ti,ab. | 0 | Advanced |
| 69 | pseudoophthalmoplegia*.ti,ab. | 0 | Advanced |
| 70 | skew deviation*.ti,ab. | 0 | Advanced |
| 71 | smooth pursuit deficienc*.ti,ab. | 0 | Advanced |
| 72 | spasm of conjugate gaze.ti,ab. | 0 | Advanced |
| 73 | tendon sheath syndrome of brown.ti,ab. | 0 | Advanced |
| 74 | fisher syndrome.ti,ab. | 1 | Advanced |
| 75 | miller fisher.ti,ab. | 0 | Advanced |
| 76 | (ophthalmoplegia, ataxia and areflexia syndrome).ti,ab. | 0 | Advanced |
| 77 | nystagmus.ti,ab. | 8 | Advanced |
| 78 | involuntary eye movement*.ti,ab. | 0 | Advanced |
| 79 | Jerky eye movement*.ti,ab. | 0 | Advanced |
| 80 | cranial nerve iii disease*.ti,ab. | 0 | Advanced |
| 81 | oculomotor nerve disease*.ti,ab. | 0 | Advanced |
| 82 | oculomotor nerve disorder*.ti,ab. | 0 | Advanced |
| 83 | oculomotor nerve pals*.ti,ab. | 0 | Advanced |
| 84 | oculomotor nerve paralys?s.ti,ab. | 0 | Advanced |
| 85 | oculomotor neuropath*.ti,ab. | 0 | Advanced |
| 86 | third cranial nerve disease*.ti,ab. | 0 | Advanced |
| 87 | third nerve pals*.ti,ab. | 0 | Advanced |
| 88 | third nerve paralysis.ti,ab. | 1 | Advanced |
| 89 | ophthalmoplegia*.ti,ab. | 1 | Advanced |
| 90 | oculomotor paralysis.ti,ab. | 0 | Advanced |
| 91 | ophthalmopares?s.ti,ab. | 0 | Advanced |
| 92 | dancing eyes.ti,ab. | 0 | Advanced |
| 93 | myoclonic encephalopathy*.ti,ab. | 0 | Advanced |
| 94 | kinsbourne syndrome.ti,ab. | 0 | Advanced |
| 95 | opsoclonus myoclonus.ti,ab. | 0 | Advanced |
| 96 | strabismus.ti,ab. | 14 | Advanced |
| 97 | dissociated horizontal deviation*.ti,ab. | 0 | Advanced |
| 98 | dissociated vertical deviation*.ti,ab. | 2 | Advanced |
| 99 | heterophoria*.ti,ab. | 0 | Advanced |
| 100 | heterotropia*.ti,ab. | 0 | Advanced |
| 101 | hypertropia*.ti,ab. | 1 | Advanced |
| 102 | phoria*.ti,ab. | 0 | Advanced |
| 103 | squint*.ti,ab. | 3 | Advanced |
| 104 | crossed eye*.ti,ab. | 0 | Advanced |
| 105 | tolosa hunt syndrome*.ti,ab. | 0 | Advanced |
| 106 | Ametropia*.ti,ab. | 0 | Advanced |
| 107 | refractive disorder*.ti,ab. | 0 | Advanced |
| 108 | refractive error*.ti,ab. | 18 | Advanced |
| 109 | aniseikonia.ti,ab. | 0 | Advanced |
| 110 | anisometropia.ti,ab. | 1 | Advanced |
| 111 | astigmatism*.ti,ab. | 12 | Advanced |
| 112 | corneal wavefront aberration*.ti,ab. | 0 | Advanced |
| 113 | farsighted*.ti,ab. | 0 | Advanced |
| 114 | longsighted*.ti,ab. | 0 | Advanced |
| 115 | hypermetropia*.ti,ab. | 1 | Advanced |
| 116 | hyperopia*.ti,ab. | 3 | Advanced |
| 117 | myopia*.ti,ab. | 14 | Advanced |
| 118 | nearsighted*.ti,ab. | 1 | Advanced |
| 119 | shortsighted*.ti,ab. | 0 | Advanced |
| 120 | presbyopia*.ti,ab. | 3 | Advanced |
| 121 | (retinal adj2 disease*).ti,ab. | 1 | Advanced |
| 122 | (cone adj2 dystroph*).ti,ab. | 0 | Advanced |
| 123 | diabetic retinopath*.ti,ab. | 29 | Advanced |
| 124 | hypertensive retinopath*.ti,ab. | 0 | Advanced |
| 125 | diabetic eye disease*.ti,ab. | 0 | Advanced |
| 126 | diabetic macular edema.ti,ab. | 2 | Advanced |
| 127 | blindness.ti,ab. | 71 | Advanced |
| 128 | hemeralopia*.ti,ab. | 0 | Advanced |
| 129 | macropsia*.ti,ab. | 0 | Advanced |
| 130 | metamorphopsia*.ti,ab. | 1 | Advanced |
| 131 | micropsia*.ti,ab. | 0 | Advanced |
| 132 | vision disabilit*.ti,ab. | 0 | Advanced |
| 133 | vision disorder*.ti,ab. | 1 | Advanced |
| 134 | vision impairment*.ti,ab. | 3 | Advanced |
| 135 | visual disorder*.ti,ab. | 0 | Advanced |
| 136 | visual impairment*.ti,ab. | 26 | Advanced |
| 137 | visual disabilit*.ti,ab. | 3 | Advanced |
| 138 | amblyopia*.ti,ab. | 14 | Advanced |
| 139 | lazy eye*.ti,ab. | 0 | Advanced |
| 140 | amauros?s.ti,ab. | 1 | Advanced |
| 141 | sudden visual loss*.ti,ab. | 0 | Advanced |
| 142 | achromatopsia*.ti,ab. | 0 | Advanced |
| 143 | colo?r vision defect*.ti,ab. | 0 | Advanced |
| 144 | colo?r vision deficienc*.ti,ab. | 0 | Advanced |
| 145 | colo?r vision impairment*.ti,ab. | 0 | Advanced |
| 146 | colo?r perception deficienc*.ti,ab. | 0 | Advanced |
| 147 | deutan defect.ti,ab. | 0 | Advanced |
| 148 | monochromatopsia.ti,ab. | 0 | Advanced |
| 149 | protan defect.ti,ab. | 0 | Advanced |
| 150 | tritan defect.ti,ab. | 0 | Advanced |
| 151 | diplopia*.ti,ab. | 13 | Advanced |
| 152 | double vision.ti,ab. | 5 | Advanced |
| 153 | polyopsia*.ti,ab. | 0 | Advanced |
| 154 | nyctalopia.ti,ab. | 1 | Advanced |
| 155 | light sensitivit*.ti,ab. | 1 | Advanced |
| 156 | photophobia*.ti,ab. | 9 | Advanced |
| 157 | scotoma*.ti,ab. | 1 | Advanced |
| 158 | retinocochleocerebral vasculopath*.ti,ab. | 0 | Advanced |
| 159 | susac* syndrome.ti,ab. | 0 | Advanced |
| 160 | diminished vision.ti,ab. | 0 | Advanced |
| 161 | reduced vision.ti,ab. | 0 | Advanced |
| 162 | subnormal vision.ti,ab. | 0 | Advanced |
| 163 | sub-normal vision.ti,ab. | 0 | Advanced |
| 164 | low vision.ti,ab. | 7 | Advanced |
| 165 | macular degeneration*.ti,ab. | 32 | Advanced |
| 166 | maculopath*.ti,ab. | 0 | Advanced |
| 167 | macular dystroph*.ti,ab. | 0 | Advanced |
| 168 | macular disorder*.ti,ab. | 0 | Advanced |
| 169 | retinal degeneration*.ti,ab. | 0 | Advanced |
| 170 | retinal degenerative disease*.ti,ab. | 0 | Advanced |
| 171 | visual field*.ti,ab. | 36 | Advanced |
| 172 | vision test*.ti,ab. | 2 | Advanced |
| 173 | colo?r perception test*.ti,ab. | 0 | Advanced |
| 174 | ocular refraction*.ti,ab. | 0 | Advanced |
| 175 | vision screening.ti,ab. | 5 | Advanced |
| 176 | visual acuit*.ti,ab. | 137 | Advanced |
| 177 | visual contrast sensitivity*.ti,ab. | 0 | Advanced |
| 178 | emmetropia*.ti,ab. | 0 | Advanced |
| 179 | automated perimetry exam*.ti,ab. | 0 | Advanced |
| 180 | campimetr*.ti,ab. | 0 | Advanced |
| 181 | perimetr*.ti,ab. | 1 | Advanced |
| 182 | tangent screen exam*.ti,ab. | 0 | Advanced |
| 183 | auditory perception.ti,ab. | 0 | Advanced |
| 184 | auditory processing.ti,ab. | 1 | Advanced |
| 185 | (auditory adj2 localization*).ti,ab. | 0 | Advanced |
| 186 | (sound adj2 localization*).ti,ab. | 0 | Advanced |
| 187 | spatial hearing.ti,ab. | 0 | Advanced |
| 188 | or/1-187 | 323 | Advanced |
| 189 | (virtual adj3 realit*).ti,ab. | 18 | Advanced |
| 190 | (augmented adj3 realit*).ti,ab. | 0 | Advanced |
| 191 | VRET.ti,ab. | 0 | Advanced |
| 192 | iVR.ti,ab. | 1 | Advanced |
| 193 | iVRs.ti,ab. | 0 | Advanced |
| 194 | iVE.ti,ab. | 0 | Advanced |
| 195 | iVEs.ti,ab. | 0 | Advanced |
| 196 | (immers* adj2 environment*).ti,ab. | 0 | Advanced |
| 197 | samsung gear.ti,ab. | 0 | Advanced |
| 198 | oculus rift.ti,ab. | 0 | Advanced |
| 199 | HCD vive.ti,ab. | 0 | Advanced |
| 200 | hololens.ti,ab. | 0 | Advanced |
| 201 | gear VR.ti,ab. | 0 | Advanced |
| 202 | head-mount*.ti,ab. | 1 | Advanced |
| 203 | (head adj3 gear*).ti,ab. | 0 | Advanced |
| 204 | HMD.ti,ab. | 1 | Advanced |
| 205 | Gen2-VR.ti,ab. | 0 | Advanced |
| 206 | (google adj2 daydream).ti,ab. | 0 | Advanced |
| 207 | (google adj2 cardboard).ti,ab. | 0 | Advanced |
| 208 | PlayStation.ti,ab. | 0 | Advanced |
| 209 | (ReTrak adj2 Utopia).ti,ab. | 0 | Advanced |
| 210 | NOON VR.ti,ab. | 0 | Advanced |
| 211 | Freefly VR.ti,ab. | 0 | Advanced |
| 212 | Homido.ti,ab. | 0 | Advanced |
| 213 | iWear.ti,ab. | 0 | Advanced |
| 214 | (mixed adj2 realit*).ti,ab. | 0 | Advanced |
| 215 | (simulat* adj2 environment*).ti,ab. | 2 | Advanced |
| 216 | (virtual adj2 environment*).ti,ab. | 3 | Advanced |
| 217 | (computer adj2 game*).ti,ab. | 6 | Advanced |
| 218 | (video adj2 game*).ti,ab. | 2 | Advanced |
| 219 | gaming.ti,ab. | 7 | Advanced |
| 220 | (user adj2 interface*).ti,ab. | 1 | Advanced |
| 221 | (virtual adj2 interface*).ti,ab. | 0 | Advanced |
| 222 | Oculus Quest.ti,ab. | 0 | Advanced |
| 223 | Meta Quest.ti,ab. | 0 | Advanced |
| 224 | Metaverse.ti,ab. | 0 | Advanced |
| 225 | (simulate* adj2 realit*).ti,ab. | 0 | Advanced |
| 226 | (tethered adj2 headset*).ti,ab. | 0 | Advanced |
| 227 | Serious game.ti,ab. | 0 | Advanced |
| 228 | or/189-227 | 30 | Advanced |
| 229 | 188 and 228 | 3 | Advanced |

Searches as run on August 16, 2024

Ovid MEDLINE(R) ALL <1946 to August 15, 2024>

| **#** | **Searches** | **Results** | **Type** |
| --- | --- | --- | --- |
| 1 | exp eye diseases/ | 657720 | Advanced |
| 2 | Optic Atrophy/ge | 782 | Advanced |
| 3 | exp Optic Nerve/ab | 2023 | Advanced |
| 4 | Visual Fields/ | 33044 | Advanced |
| 5 | exp Vision Tests/ | 121166 | Advanced |
| 6 | exp Auditory Perception/ | 87129 | Advanced |
| 7 | Sound Localization/ | 4911 | Advanced |
| 8 | (eye adj2 disease*).mp. | 56580 | Advanced |
| 9 | (eye adj2 disorder*).mp. | 2376 | Advanced |
| 10 | Albinism.mp. | 4571 | Advanced |
| 11 | (Chediak adj2 higashi syndrome).mp. | 1226 | Advanced |
| 12 | (Congenital adj2 amauros?s).mp. | 1645 | Advanced |
| 13 | dysgenesis neuroepithelialis retinae.mp. | 1 | Advanced |
| 14 | hereditary epithelial dysplasia of retina.mp. | 0 | Advanced |
| 15 | hereditary retinal aplasia.mp. | 0 | Advanced |
| 16 | heredoretinopathia congenitalis.mp. | 0 | Advanced |
| 17 | leber abiotroph*.mp. | 0 | Advanced |
| 18 | (leber* adj2 amauros?s).mp. | 1655 | Advanced |
| 19 | leber congenital tapetoretinal degeneration.mp. | 0 | Advanced |
| 20 | hereditary optic atroph*.mp. | 129 | Advanced |
| 21 | (optic adj2 hypoplasia*).mp. | 878 | Advanced |
| 22 | (retinal adj2 degeneration*).mp. | 17408 | Advanced |
| 23 | (retinal adj2 dysplasia*).mp. | 494 | Advanced |
| 24 | pigmentary retinopathy*.mp. | 628 | Advanced |
| 25 | retinitis pigmentosa.mp. | 13020 | Advanced |
| 26 | tapetoretinal degeneration*.mp. | 221 | Advanced |
| 27 | fundus flavimaculatus.mp. | 238 | Advanced |
| 28 | macular dystrophy with flecks.mp. | 2 | Advanced |
| 29 | stargardt disease.mp. | 1153 | Advanced |
| 30 | cod md syndrome*.mp. | 2 | Advanced |
| 31 | cerebromuscular dystrophy.mp. | 12 | Advanced |
| 32 | cerebroocular dysplasia muscular dystrophy syndrome.mp. | 0 | Advanced |
| 33 | chemke syndrome.mp. | 1 | Advanced |
| 34 | congenital muscular dystrophy dystroglycanopathy.mp. | 6 | Advanced |
| 35 | walker-warburg syndrome*.mp. | 475 | Advanced |
| 36 | fukuyama cmd.mp. | 27 | Advanced |
| 37 | (fukuyama adj3 muscular dystrophy).mp. | 412 | Advanced |
| 38 | fukuyama syndrome.mp. | 1 | Advanced |
| 39 | hard syndrome*.mp. | 4 | Advanced |
| 40 | lgmd2k.mp. | 7 | Advanced |
| 41 | mddga1.mp. | 0 | Advanced |
| 42 | meb syndrome.mp. | 0 | Advanced |
| 43 | muscle-eye-brain syndrome.mp. | 3 | Advanced |
| 44 | muscle eye brain disease*.mp. | 193 | Advanced |
| 45 | muscular dystrophy due to defective glycosylation of dystroglycan 4a.mp. | 0 | Advanced |
| 46 | muscular dystrophy-dystroglycanopathy.mp. | 30 | Advanced |
| 47 | pagon syndrome*.mp. | 0 | Advanced |
| 48 | warburg syndrome.mp. | 492 | Advanced |
| 49 | alpha dystroglycanopathies.mp. | 48 | Advanced |
| 50 | congenital mesodermal dysmorphodystroph*.mp. | 0 | Advanced |
| 51 | gems.mp. | 1354 | Advanced |
| 52 | glaucoma-lens ectopia-microspherophakia-stiffness-shortness syndrome.mp. | 1 | Advanced |
| 53 | (marchesani adj2 syndrome*).mp. | 216 | Advanced |
| 54 | spherophakia brachymorphia syndrome*.mp. | 5 | Advanced |
| 55 | cataract*.mp. | 81133 | Advanced |
| 56 | lens opacity*.mp. | 1134 | Advanced |
| 57 | pseudoaphakia*.mp. | 8 | Advanced |
| 58 | (lens adj2 cloud*).mp. | 69 | Advanced |
| 59 | glaucoma*.mp. | 85824 | Advanced |
| 60 | increased intraocular pressure.mp. | 1529 | Advanced |
| 61 | ocular hypertension.mp. | 10266 | Advanced |
| 62 | optic neuropathy.mp. | 12415 | Advanced |
| 63 | brown* tendon sheath syndrome*.mp. | 4 | Advanced |
| 64 | conjugate gaze spasm*.mp. | 0 | Advanced |
| 65 | convergence excess*.mp. | 119 | Advanced |
| 66 | convergence insufficienc*.mp. | 590 | Advanced |
| 67 | cyclophoria*.mp. | 24 | Advanced |
| 68 | eye motility disorder*.mp. | 19 | Advanced |
| 69 | eye movement disorder*.mp. | 476 | Advanced |
| 70 | internuclear ophthalmoplegia*.mp. | 754 | Advanced |
| 71 | ocular motility disorder*.mp. | 4564 | Advanced |
| 72 | ocular torticollis.mp. | 120 | Advanced |
| 73 | opsoclonus.mp. | 1165 | Advanced |
| 74 | parinaud* syndrome*.mp. | 341 | Advanced |
| 75 | paroxysmal ocular dyskinesia*.mp. | 0 | Advanced |
| 76 | pseudoophthalmoplegia*.mp. | 0 | Advanced |
| 77 | skew deviation*.mp. | 334 | Advanced |
| 78 | smooth pursuit deficienc*.mp. | 2 | Advanced |
| 79 | spasm of conjugate gaze.mp. | 0 | Advanced |
| 80 | tendon sheath syndrome of brown.mp. | 5 | Advanced |
| 81 | fisher syndrome.mp. | 1403 | Advanced |
| 82 | miller fisher.mp. | 1401 | Advanced |
| 83 | (ophthalmoplegia, ataxia and areflexia syndrome).mp. | 5 | Advanced |
| 84 | nystagmus.mp. | 19933 | Advanced |
| 85 | involuntary eye movement*.mp. | 171 | Advanced |
| 86 | Jerky eye movement*.mp. | 11 | Advanced |
| 87 | cranial nerve iii disease*.mp. | 0 | Advanced |
| 88 | oculomotor nerve disease*.mp. | 1884 | Advanced |
| 89 | oculomotor nerve disorder*.mp. | 2 | Advanced |
| 90 | oculomotor nerve pals*.mp. | 1035 | Advanced |
| 91 | oculomotor nerve paralys?s.mp. | 80 | Advanced |
| 92 | oculomotor neuropath*.mp. | 27 | Advanced |
| 93 | third cranial nerve disease*.mp. | 0 | Advanced |
| 94 | third nerve pals*.mp. | 835 | Advanced |
| 95 | third nerve paralysis.mp. | 39 | Advanced |
| 96 | ophthalmoplegia*.mp. | 12605 | Advanced |
| 97 | oculomotor paralysis.mp. | 1032 | Advanced |
| 98 | ophthalmopares?s.mp. | 579 | Advanced |
| 99 | dancing eyes.mp. | 41 | Advanced |
| 100 | myoclonic encephalopathy*.mp. | 273 | Advanced |
| 101 | kinsbourne syndrome.mp. | 23 | Advanced |
| 102 | opsoclonus myoclonus.mp. | 819 | Advanced |
| 103 | strabismus.mp. | 20979 | Advanced |
| 104 | dissociated horizontal deviation*.mp. | 28 | Advanced |
| 105 | dissociated vertical deviation*.mp. | 314 | Advanced |
| 106 | heterophoria*.mp. | 570 | Advanced |
| 107 | heterotropia*.mp. | 143 | Advanced |
| 108 | hypertropia*.mp. | 546 | Advanced |
| 109 | phoria*.mp. | 640 | Advanced |
| 110 | squint*.mp. | 2101 | Advanced |
| 111 | crossed eye*.mp. | 52 | Advanced |
| 112 | tolosa hunt syndrome*.mp. | 598 | Advanced |
| 113 | Ametropia*.mp. | 1118 | Advanced |
| 114 | refractive disorder*.mp. | 92 | Advanced |
| 115 | refractive error*.mp. | 17316 | Advanced |
| 116 | aniseikonia.mp. | 563 | Advanced |
| 117 | anisometropia.mp. | 2333 | Advanced |
| 118 | astigmatism*.mp. | 14836 | Advanced |
| 119 | corneal wavefront aberration*.mp. | 1029 | Advanced |
| 120 | farsighted*.mp. | 146 | Advanced |
| 121 | longsighted*.mp. | 3 | Advanced |
| 122 | hypermetropia*.mp. | 766 | Advanced |
| 123 | hyperopia*.mp. | 5673 | Advanced |
| 124 | myopia*.mp. | 30196 | Advanced |
| 125 | nearsighted*.mp. | 185 | Advanced |
| 126 | shortsighted*.mp. | 158 | Advanced |
| 127 | presbyopia*.mp. | 2805 | Advanced |
| 128 | (retinal adj2 disease*).mp. | 33260 | Advanced |
| 129 | (cone adj2 dystroph*).mp. | 1785 | Advanced |
| 130 | diabetic retinopath*.mp. | 43148 | Advanced |
| 131 | hypertensive retinopath*.mp. | 946 | Advanced |
| 132 | diabetic eye disease*.mp. | 573 | Advanced |
| 133 | diabetic macular edema.mp. | 5173 | Advanced |
| 134 | blindness.mp. | 50968 | Advanced |
| 135 | hemeralopia*.mp. | 142 | Advanced |
| 136 | macropsia*.mp. | 59 | Advanced |
| 137 | metamorphopsia*.mp. | 1044 | Advanced |
| 138 | micropsia*.mp. | 111 | Advanced |
| 139 | vision disabilit*.mp. | 83 | Advanced |
| 140 | vision disorder*.mp. | 31847 | Advanced |
| 141 | vision impairment*.mp. | 3330 | Advanced |
| 142 | visual disorder*.mp. | 1151 | Advanced |
| 143 | visual impairment*.mp. | 16645 | Advanced |
| 144 | visual disabilit*.mp. | 1124 | Advanced |
| 145 | amblyopia*.mp. | 10744 | Advanced |
| 146 | lazy eye*.mp. | 70 | Advanced |
| 147 | amauros?s.mp. | 3647 | Advanced |
| 148 | sudden visual loss*.mp. | 297 | Advanced |
| 149 | achromatopsia*.mp. | 698 | Advanced |
| 150 | colo?r vision defect*.mp. | 4443 | Advanced |
| 151 | colo?r vision deficienc*.mp. | 570 | Advanced |
| 152 | colo?r vision impairment*.mp. | 95 | Advanced |
| 153 | colo?r perception deficienc*.mp. | 3 | Advanced |
| 154 | deutan defect.mp. | 7 | Advanced |
| 155 | monochromatopsia.mp. | 0 | Advanced |
| 156 | protan defect.mp. | 7 | Advanced |
| 157 | tritan defect.mp. | 22 | Advanced |
| 158 | diplopia*.mp. | 13434 | Advanced |
| 159 | double vision.mp. | 1482 | Advanced |
| 160 | polyopsia*.mp. | 3 | Advanced |
| 161 | nyctalopia.mp. | 471 | Advanced |
| 162 | light sensitivit*.mp. | 2227 | Advanced |
| 163 | photophobia*.mp. | 4506 | Advanced |
| 164 | scotoma*.mp. | 5733 | Advanced |
| 165 | retinocochleocerebral vasculopath*.mp. | 16 | Advanced |
| 166 | susac* syndrome.mp. | 515 | Advanced |
| 167 | diminished vision.mp. | 167 | Advanced |
| 168 | reduced vision.mp. | 944 | Advanced |
| 169 | subnormal vision.mp. | 99 | Advanced |
| 170 | sub-normal vision.mp. | 9 | Advanced |
| 171 | low vision.mp. | 3671 | Advanced |
| 172 | macular degeneration*.mp. | 33240 | Advanced |
| 173 | maculopath*.mp. | 5982 | Advanced |
| 174 | macular dystroph*.mp. | 1953 | Advanced |
| 175 | macular disorder*.mp. | 187 | Advanced |
| 176 | retinal degeneration*.mp. | 16388 | Advanced |
| 177 | retinal degenerative disease*.mp. | 1244 | Advanced |
| 178 | visual field*.mp. | 53523 | Advanced |
| 179 | vision test*.mp. | 12194 | Advanced |
| 180 | colo?r perception test*.mp. | 2618 | Advanced |
| 181 | ocular refraction*.mp. | 252 | Advanced |
| 182 | vision screening.mp. | 3224 | Advanced |
| 183 | visual acuit*.mp. | 126951 | Advanced |
| 184 | visual contrast sensitivity*.mp. | 235 | Advanced |
| 185 | emmetropia*.mp. | 2036 | Advanced |
| 186 | automated perimetry exam*.mp. | 9 | Advanced |
| 187 | campimetr*.mp. | 322 | Advanced |
| 188 | perimetr*.mp. | 8502 | Advanced |
| 189 | tangent screen exam*.mp. | 13 | Advanced |
| 190 | auditory perception.mp. | 33898 | Advanced |
| 191 | auditory processing.mp. | 5434 | Advanced |
| 192 | (auditory adj2 localization*).mp. | 488 | Advanced |
| 193 | (sound adj2 localization*).mp. | 6026 | Advanced |
| 194 | spatial hearing.mp. | 569 | Advanced |
| 195 | or/1-194 | 914767 | Advanced |
| 196 | Virtual Reality/ | 6869 | Advanced |
| 197 | (virtual adj3 realit*).mp. | 22024 | Advanced |
| 198 | (augmented adj3 realit*).mp. | 5913 | Advanced |
| 199 | VRET.mp. | 156 | Advanced |
| 200 | iVR.mp. | 2057 | Advanced |
| 201 | iVRs.mp. | 220 | Advanced |
| 202 | iVE.mp. | 1951 | Advanced |
| 203 | iVEs.mp. | 195 | Advanced |
| 204 | (immers* adj2 environment*).mp. | 918 | Advanced |
| 205 | samsung gear.mp. | 42 | Advanced |
| 206 | oculus rift.mp. | 132 | Advanced |
| 207 | HCD vive.mp. | 0 | Advanced |
| 208 | hololens.mp. | 439 | Advanced |
| 209 | gear VR.mp. | 16 | Advanced |
| 210 | head-mount*.mp. | 2934 | Advanced |
| 211 | (head adj3 gear*).mp. | 101 | Advanced |
| 212 | HMD.mp. | 1497 | Advanced |
| 213 | Gen2-VR.mp. | 1 | Advanced |
| 214 | (google adj2 daydream).mp. | 5 | Advanced |
| 215 | (google adj2 cardboard).mp. | 23 | Advanced |
| 216 | PlayStation.mp. | 91 | Advanced |
| 217 | (ReTrak adj2 Utopia).mp. | 0 | Advanced |
| 218 | NOON VR.mp. | 0 | Advanced |
| 219 | Freefly VR.mp. | 0 | Advanced |
| 220 | Homido.mp. | 0 | Advanced |
| 221 | iWear.mp. | 0 | Advanced |
| 222 | (mixed adj2 realit*).mp. | 1297 | Advanced |
| 223 | (simulat* adj2 environment*).mp. | 6537 | Advanced |
| 224 | (virtual adj2 environment*).mp. | 6845 | Advanced |
| 225 | Video Games/ | 7667 | Advanced |
| 226 | (computer adj2 game*).mp. | 1875 | Advanced |
| 227 | (video adj2 game*).mp. | 10420 | Advanced |
| 228 | gaming.mp. | 6266 | Advanced |
| 229 | user-computer interface/ | 39824 | Advanced |
| 230 | (user adj2 interface*).mp. | 48401 | Advanced |
| 231 | (virtual adj2 interface*).mp. | 214 | Advanced |
| 232 | Oculus Quest.mp. | 46 | Advanced |
| 233 | Meta Quest.mp. | 25 | Advanced |
| 234 | Metaverse.mp. | 547 | Advanced |
| 235 | (simulate* adj2 realit*).mp. | 126 | Advanced |
| 236 | (tethered adj2 headset*).mp. | 1 | Advanced |
| 237 | Serious game.mp. | 903 | Advanced |
| 238 | or/196-237 | 98233 | Advanced |
| 239 | 195 and 238 | 3109 | Advanced |
| 240 | animals/ not (animals/ and humans/) | 5214906 | Advanced |
| 241 | 239 not 240 | 3038 | Advanced |
| 242 | limit 241 to english language | 2940 | Advanced |
| 243 | remove duplicates from 242 | 2935 | Advanced |
| 244 | limit 243 to dt="20230814-20240816" | 212 | Advanced |

Embase <1974 to 2024 August 15>

| **#** | **Searches** | **Results** | **Type** |
| --- | --- | --- | --- |
| 1 | exp eye disease/ | 1144906 | Advanced |
| 2 | exp hereditary optic atrophy/ | 7889 | Advanced |
| 3 | visual field/ | 40865 | Advanced |
| 4 | exp vision test/ | 46013 | Advanced |
| 5 | exp hearing/ | 72047 | Advanced |
| 6 | sound detection/ | 11793 | Advanced |
| 7 | (eye adj2 disease*).mp. | 70213 | Advanced |
| 8 | (eye adj2 disorder*).mp. | 9150 | Advanced |
| 9 | Albinism.mp. | 7105 | Advanced |
| 10 | (Chediak adj2 higashi syndrome).mp. | 1783 | Advanced |
| 11 | (Congenital adj2 amauros?s).mp. | 2993 | Advanced |
| 12 | dysgenesis neuroepithelialis retinae.mp. | 0 | Advanced |
| 13 | hereditary epithelial dysplasia of retina.mp. | 0 | Advanced |
| 14 | hereditary retinal aplasia.mp. | 0 | Advanced |
| 15 | heredoretinopathia congenitalis.mp. | 0 | Advanced |
| 16 | leber abiotroph*.mp. | 0 | Advanced |
| 17 | (leber* adj2 amauros?s).mp. | 3001 | Advanced |
| 18 | leber congenital tapetoretinal degeneration.mp. | 0 | Advanced |
| 19 | hereditary optic atroph*.mp. | 1059 | Advanced |
| 20 | (optic adj2 hypoplasia*).mp. | 1754 | Advanced |
| 21 | (retinal adj2 degeneration*).mp. | 15280 | Advanced |
| 22 | (retinal adj2 dysplasia*).mp. | 442 | Advanced |
| 23 | pigmentary retinopathy*.mp. | 851 | Advanced |
| 24 | retinitis pigmentosa.mp. | 17401 | Advanced |
| 25 | tapetoretinal degeneration*.mp. | 612 | Advanced |
| 26 | fundus flavimaculatus.mp. | 283 | Advanced |
| 27 | macular dystrophy with flecks.mp. | 2 | Advanced |
| 28 | stargardt disease.mp. | 2607 | Advanced |
| 29 | cod md syndrome*.mp. | 3 | Advanced |
| 30 | cerebromuscular dystrophy.mp. | 14 | Advanced |
| 31 | cerebroocular dysplasia muscular dystrophy syndrome.mp. | 0 | Advanced |
| 32 | chemke syndrome.mp. | 1 | Advanced |
| 33 | congenital muscular dystrophy dystroglycanopathy.mp. | 13 | Advanced |
| 34 | walker-warburg syndrome*.mp. | 894 | Advanced |
| 35 | fukuyama cmd.mp. | 42 | Advanced |
| 36 | (fukuyama adj3 muscular dystrophy).mp. | 714 | Advanced |
| 37 | fukuyama syndrome.mp. | 2 | Advanced |
| 38 | hard syndrome*.mp. | 5 | Advanced |
| 39 | lgmd2k.mp. | 11 | Advanced |
| 40 | mddga1.mp. | 2 | Advanced |
| 41 | meb syndrome.mp. | 0 | Advanced |
| 42 | muscle-eye-brain syndrome.mp. | 3 | Advanced |
| 43 | muscle eye brain disease*.mp. | 405 | Advanced |
| 44 | muscular dystrophy due to defective glycosylation of dystroglycan 4a.mp. | 0 | Advanced |
| 45 | muscular dystrophy-dystroglycanopathy.mp. | 56 | Advanced |
| 46 | pagon syndrome*.mp. | 0 | Advanced |
| 47 | warburg syndrome.mp. | 903 | Advanced |
| 48 | alpha dystroglycanopathies.mp. | 92 | Advanced |
| 49 | congenital mesodermal dysmorphodystroph*.mp. | 0 | Advanced |
| 50 | gems.mp. | 2039 | Advanced |
| 51 | glaucoma-lens ectopia-microspherophakia-stiffness-shortness syndrome.mp. | 1 | Advanced |
| 52 | (marchesani adj2 syndrome*).mp. | 326 | Advanced |
| 53 | spherophakia brachymorphia syndrome*.mp. | 2 | Advanced |
| 54 | cataract*.mp. | 114907 | Advanced |
| 55 | lens opacity*.mp. | 1451 | Advanced |
| 56 | pseudoaphakia*.mp. | 11 | Advanced |
| 57 | (lens adj2 cloud*).mp. | 93 | Advanced |
| 58 | glaucoma*.mp. | 115477 | Advanced |
| 59 | increased intraocular pressure.mp. | 1847 | Advanced |
| 60 | ocular hypertension.mp. | 8306 | Advanced |
| 61 | optic neuropathy.mp. | 19547 | Advanced |
| 62 | brown* tendon sheath syndrome*.mp. | 3 | Advanced |
| 63 | conjugate gaze spasm*.mp. | 0 | Advanced |
| 64 | convergence excess*.mp. | 129 | Advanced |
| 65 | convergence insufficienc*.mp. | 716 | Advanced |
| 66 | cyclophoria*.mp. | 23 | Advanced |
| 67 | eye motility disorder*.mp. | 25 | Advanced |
| 68 | eye movement disorder*.mp. | 6588 | Advanced |
| 69 | internuclear ophthalmoplegia*.mp. | 1453 | Advanced |
| 70 | ocular motility disorder*.mp. | 540 | Advanced |
| 71 | ocular torticollis.mp. | 98 | Advanced |
| 72 | opsoclonus.mp. | 2153 | Advanced |
| 73 | parinaud* syndrome*.mp. | 478 | Advanced |
| 74 | paroxysmal ocular dyskinesia*.mp. | 0 | Advanced |
| 75 | pseudoophthalmoplegia*.mp. | 1 | Advanced |
| 76 | skew deviation*.mp. | 529 | Advanced |
| 77 | smooth pursuit deficienc*.mp. | 2 | Advanced |
| 78 | spasm of conjugate gaze.mp. | 1 | Advanced |
| 79 | tendon sheath syndrome of brown.mp. | 4 | Advanced |
| 80 | fisher syndrome.mp. | 1852 | Advanced |
| 81 | miller fisher.mp. | 1729 | Advanced |
| 82 | (ophthalmoplegia, ataxia and areflexia syndrome).mp. | 5 | Advanced |
| 83 | nystagmus.mp. | 32029 | Advanced |
| 84 | involuntary eye movement*.mp. | 203 | Advanced |
| 85 | Jerky eye movement*.mp. | 29 | Advanced |
| 86 | cranial nerve iii disease*.mp. | 0 | Advanced |
| 87 | oculomotor nerve disease*.mp. | 1427 | Advanced |
| 88 | oculomotor nerve disorder*.mp. | 4 | Advanced |
| 89 | oculomotor nerve pals*.mp. | 1382 | Advanced |
| 90 | oculomotor nerve paralys?s.mp. | 99 | Advanced |
| 91 | oculomotor neuropath*.mp. | 33 | Advanced |
| 92 | third cranial nerve disease*.mp. | 0 | Advanced |
| 93 | third nerve pals*.mp. | 1115 | Advanced |
| 94 | third nerve paralysis.mp. | 39 | Advanced |
| 95 | ophthalmoplegia*.mp. | 19817 | Advanced |
| 96 | oculomotor paralysis.mp. | 220 | Advanced |
| 97 | ophthalmopares?s.mp. | 950 | Advanced |
| 98 | dancing eyes.mp. | 63 | Advanced |
| 99 | myoclonic encephalopathy*.mp. | 419 | Advanced |
| 100 | kinsbourne syndrome.mp. | 60 | Advanced |
| 101 | opsoclonus myoclonus.mp. | 1472 | Advanced |
| 102 | strabismus.mp. | 33509 | Advanced |
| 103 | dissociated horizontal deviation*.mp. | 37 | Advanced |
| 104 | dissociated vertical deviation*.mp. | 386 | Advanced |
| 105 | heterophoria*.mp. | 541 | Advanced |
| 106 | heterotropia*.mp. | 159 | Advanced |
| 107 | hypertropia*.mp. | 671 | Advanced |
| 108 | phoria*.mp. | 691 | Advanced |
| 109 | squint*.mp. | 2229 | Advanced |
| 110 | crossed eye*.mp. | 42 | Advanced |
| 111 | tolosa hunt syndrome*.mp. | 1031 | Advanced |
| 112 | Ametropia*.mp. | 1631 | Advanced |
| 113 | refractive disorder*.mp. | 114 | Advanced |
| 114 | refractive error*.mp. | 15745 | Advanced |
| 115 | aniseikonia.mp. | 565 | Advanced |
| 116 | anisometropia.mp. | 4117 | Advanced |
| 117 | astigmatism*.mp. | 20656 | Advanced |
| 118 | corneal wavefront aberration*.mp. | 724 | Advanced |
| 119 | farsighted*.mp. | 174 | Advanced |
| 120 | longsighted*.mp. | 9 | Advanced |
| 121 | hypermetropia*.mp. | 8435 | Advanced |
| 122 | hyperopia*.mp. | 4632 | Advanced |
| 123 | myopia*.mp. | 40640 | Advanced |
| 124 | nearsighted*.mp. | 204 | Advanced |
| 125 | shortsighted*.mp. | 176 | Advanced |
| 126 | presbyopia*.mp. | 3728 | Advanced |
| 127 | (retinal adj2 disease*).mp. | 17068 | Advanced |
| 128 | (cone adj2 dystroph*).mp. | 2824 | Advanced |
| 129 | diabetic retinopath*.mp. | 66862 | Advanced |
| 130 | hypertensive retinopath*.mp. | 1259 | Advanced |
| 131 | diabetic eye disease*.mp. | 1116 | Advanced |
| 132 | diabetic macular edema.mp. | 11131 | Advanced |
| 133 | blindness.mp. | 77589 | Advanced |
| 134 | hemeralopia*.mp. | 91 | Advanced |
| 135 | macropsia*.mp. | 129 | Advanced |
| 136 | metamorphopsia*.mp. | 1846 | Advanced |
| 137 | micropsia*.mp. | 209 | Advanced |
| 138 | vision disabilit*.mp. | 107 | Advanced |
| 139 | vision disorder*.mp. | 1672 | Advanced |
| 140 | vision impairment*.mp. | 4423 | Advanced |
| 141 | visual disorder*.mp. | 36013 | Advanced |
| 142 | visual impairment*.mp. | 77301 | Advanced |
| 143 | visual disabilit*.mp. | 1490 | Advanced |
| 144 | amblyopia*.mp. | 14252 | Advanced |
| 145 | lazy eye*.mp. | 103 | Advanced |
| 146 | amauros?s.mp. | 5475 | Advanced |
| 147 | sudden visual loss*.mp. | 394 | Advanced |
| 148 | achromatopsia*.mp. | 979 | Advanced |
| 149 | colo?r vision defect*.mp. | 4354 | Advanced |
| 150 | colo?r vision deficienc*.mp. | 753 | Advanced |
| 151 | colo?r vision impairment*.mp. | 146 | Advanced |
| 152 | colo?r perception deficienc*.mp. | 3 | Advanced |
| 153 | deutan defect.mp. | 10 | Advanced |
| 154 | monochromatopsia.mp. | 1 | Advanced |
| 155 | protan defect.mp. | 15 | Advanced |
| 156 | tritan defect.mp. | 33 | Advanced |
| 157 | diplopia*.mp. | 34000 | Advanced |
| 158 | double vision.mp. | 2440 | Advanced |
| 159 | polyopsia*.mp. | 3 | Advanced |
| 160 | nyctalopia.mp. | 613 | Advanced |
| 161 | light sensitivit*.mp. | 2833 | Advanced |
| 162 | photophobia*.mp. | 16256 | Advanced |
| 163 | scotoma*.mp. | 9200 | Advanced |
| 164 | retinocochleocerebral vasculopath*.mp. | 21 | Advanced |
| 165 | susac* syndrome.mp. | 901 | Advanced |
| 166 | diminished vision.mp. | 220 | Advanced |
| 167 | reduced vision.mp. | 1195 | Advanced |
| 168 | subnormal vision.mp. | 90 | Advanced |
| 169 | sub-normal vision.mp. | 7 | Advanced |
| 170 | low vision.mp. | 9413 | Advanced |
| 171 | macular degeneration*.mp. | 45622 | Advanced |
| 172 | maculopath*.mp. | 12401 | Advanced |
| 173 | macular dystroph*.mp. | 2443 | Advanced |
| 174 | macular disorder*.mp. | 236 | Advanced |
| 175 | retinal degeneration*.mp. | 13667 | Advanced |
| 176 | retinal degenerative disease*.mp. | 1692 | Advanced |
| 177 | visual field*.mp. | 67628 | Advanced |
| 178 | vision test*.mp. | 12803 | Advanced |
| 179 | colo?r perception test*.mp. | 42 | Advanced |
| 180 | ocular refraction*.mp. | 278 | Advanced |
| 181 | vision screening.mp. | 1927 | Advanced |
| 182 | visual acuit*.mp. | 184594 | Advanced |
| 183 | visual contrast sensitivity*.mp. | 304 | Advanced |
| 184 | emmetropia*.mp. | 3213 | Advanced |
| 185 | automated perimetry exam*.mp. | 9 | Advanced |
| 186 | campimetr*.mp. | 401 | Advanced |
| 187 | perimetr*.mp. | 19315 | Advanced |
| 188 | tangent screen exam*.mp. | 13 | Advanced |
| 189 | auditory perception.mp. | 3275 | Advanced |
| 190 | auditory processing.mp. | 6956 | Advanced |
| 191 | (auditory adj2 localization*).mp. | 542 | Advanced |
| 192 | (sound adj2 localization*).mp. | 2913 | Advanced |
| 193 | spatial hearing.mp. | 664 | Advanced |
| 194 | or/1-193 | 1363630 | Advanced |
| 195 | virtual reality/ | 30077 | Advanced |
| 196 | augmented reality/ | 3384 | Advanced |
| 197 | (virtual adj3 realit*).mp. | 37734 | Advanced |
| 198 | (augmented adj3 realit*).mp. | 7029 | Advanced |
| 199 | VRET.mp. | 192 | Advanced |
| 200 | iVR.mp. | 2913 | Advanced |
| 201 | iVRs.mp. | 410 | Advanced |
| 202 | iVE.mp. | 3381 | Advanced |
| 203 | iVEs.mp. | 436 | Advanced |
| 204 | (immers* adj2 environment*).mp. | 1035 | Advanced |
| 205 | samsung gear.mp. | 78 | Advanced |
| 206 | oculus rift.mp. | 298 | Advanced |
| 207 | HCD vive.mp. | 0 | Advanced |
| 208 | hololens.mp. | 694 | Advanced |
| 209 | gear VR.mp. | 60 | Advanced |
| 210 | head-mount*.mp. | 4607 | Advanced |
| 211 | (head adj3 gear*).mp. | 134 | Advanced |
| 212 | HMD.mp. | 1892 | Advanced |
| 213 | Gen2-VR.mp. | 1 | Advanced |
| 214 | (google adj2 daydream).mp. | 9 | Advanced |
| 215 | (google adj2 cardboard).mp. | 44 | Advanced |
| 216 | PlayStation.mp. | 151 | Advanced |
| 217 | (ReTrak adj2 Utopia).mp. | 0 | Advanced |
| 218 | NOON VR.mp. | 0 | Advanced |
| 219 | Freefly VR.mp. | 0 | Advanced |
| 220 | Homido.mp. | 0 | Advanced |
| 221 | iWear.mp. | 5 | Advanced |
| 222 | (mixed adj2 realit*).mp. | 1416 | Advanced |
| 223 | (simulat* adj2 environment*).mp. | 7586 | Advanced |
| 224 | (virtual adj2 environment*).mp. | 9289 | Advanced |
| 225 | exp video game/ | 7365 | Advanced |
| 226 | (computer adj2 game*).mp. | 2533 | Advanced |
| 227 | (video adj2 game*).mp. | 10420 | Advanced |
| 228 | gaming.mp. | 7803 | Advanced |
| 229 | computer interface/ | 37589 | Advanced |
| 230 | (user adj2 interface*).mp. | 13981 | Advanced |
| 231 | (virtual adj2 interface*).mp. | 254 | Advanced |
| 232 | Oculus Quest.mp. | 111 | Advanced |
| 233 | Meta Quest.mp. | 41 | Advanced |
| 234 | Metaverse.mp. | 462 | Advanced |
| 235 | (simulate* adj2 realit*).mp. | 162 | Advanced |
| 236 | (tethered adj2 headset*).mp. | 2 | Advanced |
| 237 | Serious game.mp. | 867 | Advanced |
| 238 | or/195-237 | 121069 | Advanced |
| 239 | 194 and 238 | 4945 | Advanced |
| 240 | (exp animals/ or exp animal experimentation/ or nonhuman/) not ((exp animals/ or exp animal experimentation/ or nonhuman/) and exp human/) | 7514249 | Advanced |
| 241 | 239 not 240 | 4775 | Advanced |
| 242 | limit 241 to english language | 4617 | Advanced |
| 243 | limit 242 to (conference abstract or conference paper or "conference review" or conference proceeding) | 927 | Advanced |
| 244 | 242 not 243 | 3690 | Advanced |
| 245 | remove duplicates from 244 | 3605 | Advanced |
| 246 | limit 245 to dc="20230814-20240816" | 423 | Advanced |

EBM Reviews - Cochrane Central Register of Controlled Trials

| **#** | **Searches** | **Results** | **Type** |
| --- | --- | --- | --- |
| 1 | exp eye diseases/ | 26855 | Advanced |
| 2 | Optic Atrophy/ge | 0 | Advanced |
| 3 | exp Optic Nerve/ab | 0 | Advanced |
| 4 | Visual Fields/ | 1161 | Advanced |
| 5 | exp Vision Tests/ | 8200 | Advanced |
| 6 | exp Auditory Perception/ | 2603 | Advanced |
| 7 | Sound Localization/ | 113 | Advanced |
| 8 | (eye adj2 disease*).mp. | 4869 | Advanced |
| 9 | (eye adj2 disorder*).mp. | 580 | Advanced |
| 10 | Albinism.mp. | 24 | Advanced |
| 11 | (Chediak adj2 higashi syndrome).mp. | 2 | Advanced |
| 12 | (Congenital adj2 amauros?s).mp. | 32 | Advanced |
| 13 | dysgenesis neuroepithelialis retinae.mp. | 0 | Advanced |
| 14 | hereditary epithelial dysplasia of retina.mp. | 0 | Advanced |
| 15 | hereditary retinal aplasia.mp. | 0 | Advanced |
| 16 | heredoretinopathia congenitalis.mp. | 0 | Advanced |
| 17 | leber abiotroph*.mp. | 0 | Advanced |
| 18 | (leber* adj2 amauros?s).mp. | 31 | Advanced |
| 19 | leber congenital tapetoretinal degeneration.mp. | 0 | Advanced |
| 20 | hereditary optic atroph*.mp. | 2 | Advanced |
| 21 | (optic adj2 hypoplasia*).mp. | 11 | Advanced |
| 22 | (retinal adj2 degeneration*).mp. | 168 | Advanced |
| 23 | (retinal adj2 dysplasia*).mp. | 1 | Advanced |
| 24 | pigmentary retinopathy*.mp. | 5 | Advanced |
| 25 | retinitis pigmentosa.mp. | 337 | Advanced |
| 26 | tapetoretinal degeneration*.mp. | 0 | Advanced |
| 27 | fundus flavimaculatus.mp. | 6 | Advanced |
| 28 | macular dystrophy with flecks.mp. | 0 | Advanced |
| 29 | stargardt disease.mp. | 59 | Advanced |
| 30 | cod md syndrome*.mp. | 0 | Advanced |
| 31 | cerebromuscular dystrophy.mp. | 0 | Advanced |
| 32 | cerebroocular dysplasia muscular dystrophy syndrome.mp. | 0 | Advanced |
| 33 | chemke syndrome.mp. | 0 | Advanced |
| 34 | congenital muscular dystrophy dystroglycanopathy.mp. | 0 | Advanced |
| 35 | walker-warburg syndrome*.mp. | 0 | Advanced |
| 36 | fukuyama cmd.mp. | 0 | Advanced |
| 37 | (fukuyama adj3 muscular dystrophy).mp. | 8 | Advanced |
| 38 | fukuyama syndrome.mp. | 0 | Advanced |
| 39 | hard syndrome*.mp. | 0 | Advanced |
| 40 | lgmd2k.mp. | 0 | Advanced |
| 41 | mddga1.mp. | 0 | Advanced |
| 42 | meb syndrome.mp. | 0 | Advanced |
| 43 | muscle-eye-brain syndrome.mp. | 0 | Advanced |
| 44 | muscle eye brain disease*.mp. | 1 | Advanced |
| 45 | muscular dystrophy due to defective glycosylation of dystroglycan 4a.mp. | 0 | Advanced |
| 46 | muscular dystrophy-dystroglycanopathy.mp. | 0 | Advanced |
| 47 | pagon syndrome*.mp. | 0 | Advanced |
| 48 | warburg syndrome.mp. | 0 | Advanced |
| 49 | alpha dystroglycanopathies.mp. | 0 | Advanced |
| 50 | congenital mesodermal dysmorphodystroph*.mp. | 0 | Advanced |
| 51 | gems.mp. | 98 | Advanced |
| 52 | glaucoma-lens ectopia-microspherophakia-stiffness-shortness syndrome.mp. | 0 | Advanced |
| 53 | (marchesani adj2 syndrome*).mp. | 0 | Advanced |
| 54 | spherophakia brachymorphia syndrome*.mp. | 0 | Advanced |
| 55 | cataract*.mp. | 9413 | Advanced |
| 56 | lens opacity*.mp. | 125 | Advanced |
| 57 | pseudoaphakia*.mp. | 0 | Advanced |
| 58 | (lens adj2 cloud*).mp. | 13 | Advanced |
| 59 | glaucoma*.mp. | 9157 | Advanced |
| 60 | increased intraocular pressure.mp. | 170 | Advanced |
| 61 | ocular hypertension.mp. | 2783 | Advanced |
| 62 | optic neuropathy.mp. | 549 | Advanced |
| 63 | brown* tendon sheath syndrome*.mp. | 0 | Advanced |
| 64 | conjugate gaze spasm*.mp. | 0 | Advanced |
| 65 | convergence excess*.mp. | 12 | Advanced |
| 66 | convergence insufficienc*.mp. | 129 | Advanced |
| 67 | cyclophoria*.mp. | 1 | Advanced |
| 68 | eye motility disorder*.mp. | 0 | Advanced |
| 69 | eye movement disorder*.mp. | 54 | Advanced |
| 70 | internuclear ophthalmoplegia*.mp. | 10 | Advanced |
| 71 | ocular motility disorder*.mp. | 122 | Advanced |
| 72 | ocular torticollis.mp. | 1 | Advanced |
| 73 | opsoclonus.mp. | 14 | Advanced |
| 74 | parinaud* syndrome*.mp. | 1 | Advanced |
| 75 | paroxysmal ocular dyskinesia*.mp. | 0 | Advanced |
| 76 | pseudoophthalmoplegia*.mp. | 0 | Advanced |
| 77 | skew deviation*.mp. | 4 | Advanced |
| 78 | smooth pursuit deficienc*.mp. | 0 | Advanced |
| 79 | spasm of conjugate gaze.mp. | 0 | Advanced |
| 80 | tendon sheath syndrome of brown.mp. | 0 | Advanced |
| 81 | fisher syndrome.mp. | 5 | Advanced |
| 82 | miller fisher.mp. | 3 | Advanced |
| 83 | (ophthalmoplegia, ataxia and areflexia syndrome).mp. [mp=title, original title, abstract, floating sub-heading word, mesh headings, heading words, keyword] | 0 | Advanced |
| 84 | nystagmus.mp. | 713 | Advanced |
| 85 | involuntary eye movement*.mp. | 7 | Advanced |
| 86 | Jerky eye movement*.mp. | 0 | Advanced |
| 87 | cranial nerve iii disease*.mp. | 0 | Advanced |
| 88 | oculomotor nerve disease*.mp. | 13 | Advanced |
| 89 | oculomotor nerve disorder*.mp. | 0 | Advanced |
| 90 | oculomotor nerve pals*.mp. | 12 | Advanced |
| 91 | oculomotor nerve paralys?s.mp. | 0 | Advanced |
| 92 | oculomotor neuropath*.mp. | 0 | Advanced |
| 93 | third cranial nerve disease*.mp. | 0 | Advanced |
| 94 | third nerve pals*.mp. | 2 | Advanced |
| 95 | third nerve paralysis.mp. | 0 | Advanced |
| 96 | ophthalmoplegia*.mp. | 99 | Advanced |
| 97 | oculomotor paralysis.mp. | 7 | Advanced |
| 98 | ophthalmopares?s.mp. | 6 | Advanced |
| 99 | dancing eyes.mp. | 0 | Advanced |
| 100 | myoclonic encephalopathy*.mp. | 2 | Advanced |
| 101 | kinsbourne syndrome.mp. | 0 | Advanced |
| 102 | opsoclonus myoclonus.mp. | 13 | Advanced |
| 103 | strabismus.mp. | 1471 | Advanced |
| 104 | dissociated horizontal deviation*.mp. | 0 | Advanced |
| 105 | dissociated vertical deviation*.mp. | 17 | Advanced |
| 106 | heterophoria*.mp. | 63 | Advanced |
| 107 | heterotropia*.mp. | 15 | Advanced |
| 108 | hypertropia*.mp. | 15 | Advanced |
| 109 | phoria*.mp. | 136 | Advanced |
| 110 | squint*.mp. | 150 | Advanced |
| 111 | crossed eye*.mp. | 1 | Advanced |
| 112 | tolosa hunt syndrome*.mp. | 1 | Advanced |
| 113 | Ametropia*.mp. | 142 | Advanced |
| 114 | refractive disorder*.mp. | 4 | Advanced |
| 115 | refractive error*.mp. | 1937 | Advanced |
| 116 | aniseikonia.mp. | 22 | Advanced |
| 117 | anisometropia.mp. | 373 | Advanced |
| 118 | astigmatism*.mp. | 2524 | Advanced |
| 119 | corneal wavefront aberration*.mp. | 166 | Advanced |
| 120 | farsighted*.mp. | 13 | Advanced |
| 121 | longsighted*.mp. | 0 | Advanced |
| 122 | hypermetropia*.mp. | 154 | Advanced |
| 123 | hyperopia*.mp. | 336 | Advanced |
| 124 | myopia*.mp. | 3753 | Advanced |
| 125 | nearsighted*.mp. | 61 | Advanced |
| 126 | shortsighted*.mp. | 11 | Advanced |
| 127 | presbyopia*.mp. | 451 | Advanced |
| 128 | (retinal adj2 disease*).mp. | 1042 | Advanced |
| 129 | (cone adj2 dystroph*).mp. | 24 | Advanced |
| 130 | diabetic retinopath*.mp. | 4759 | Advanced |
| 131 | hypertensive retinopath*.mp. | 30 | Advanced |
| 132 | diabetic eye disease*.mp. | 96 | Advanced |
| 133 | diabetic macular edema.mp. | 1896 | Advanced |
| 134 | blindness.mp. | 2854 | Advanced |
| 135 | hemeralopia*.mp. | 0 | Advanced |
| 136 | macropsia*.mp. | 0 | Advanced |
| 137 | metamorphopsia*.mp. | 104 | Advanced |
| 138 | micropsia*.mp. | 3 | Advanced |
| 139 | vision disabilit*.mp. | 2 | Advanced |
| 140 | vision disorder*.mp. | 1040 | Advanced |
| 141 | vision impairment*.mp. | 275 | Advanced |
| 142 | visual disorder*.mp. | 706 | Advanced |
| 143 | visual impairment*.mp. | 2257 | Advanced |
| 144 | visual disabilit*.mp. | 71 | Advanced |
| 145 | amblyopia*.mp. | 881 | Advanced |
| 146 | lazy eye*.mp. | 30 | Advanced |
| 147 | amauros?s.mp. | 97 | Advanced |
| 148 | sudden visual loss*.mp. | 2 | Advanced |
| 149 | achromatopsia*.mp. | 14 | Advanced |
| 150 | colo?r vision defect*.mp. | 98 | Advanced |
| 151 | colo?r vision deficienc*.mp. | 22 | Advanced |
| 152 | colo?r vision impairment*.mp. | 8 | Advanced |
| 153 | colo?r perception deficienc*.mp. | 0 | Advanced |
| 154 | deutan defect.mp. | 1 | Advanced |
| 155 | monochromatopsia.mp. | 0 | Advanced |
| 156 | protan defect.mp. | 2 | Advanced |
| 157 | tritan defect.mp. | 1 | Advanced |
| 158 | diplopia*.mp. | 758 | Advanced |
| 159 | double vision.mp. | 106 | Advanced |
| 160 | polyopsia*.mp. | 0 | Advanced |
| 161 | nyctalopia.mp. | 4 | Advanced |
| 162 | light sensitivit*.mp. | 173 | Advanced |
| 163 | photophobia*.mp. | 1266 | Advanced |
| 164 | scotoma*.mp. | 291 | Advanced |
| 165 | retinocochleocerebral vasculopath*.mp. | 0 | Advanced |
| 166 | susac* syndrome.mp. | 2 | Advanced |
| 167 | diminished vision.mp. | 14 | Advanced |
| 168 | reduced vision.mp. | 75 | Advanced |
| 169 | subnormal vision.mp. | 3 | Advanced |
| 170 | sub-normal vision.mp. | 0 | Advanced |
| 171 | low vision.mp. | 570 | Advanced |
| 172 | macular degeneration*.mp. | 3978 | Advanced |
| 173 | maculopath*.mp. | 461 | Advanced |
| 174 | macular dystroph*.mp. | 36 | Advanced |
| 175 | macular disorder*.mp. | 9 | Advanced |
| 176 | retinal degeneration*.mp. | 119 | Advanced |
| 177 | retinal degenerative disease*.mp. | 18 | Advanced |
| 178 | visual field*.mp. | 3889 | Advanced |
| 179 | vision test*.mp. | 733 | Advanced |
| 180 | colo?r perception test*.mp. | 56 | Advanced |
| 181 | ocular refraction*.mp. | 15 | Advanced |
| 182 | vision screening.mp. | 266 | Advanced |
| 183 | visual acuit*.mp. | 18287 | Advanced |
| 184 | visual contrast sensitivity*.mp. | 31 | Advanced |
| 185 | emmetropia*.mp. | 185 | Advanced |
| 186 | automated perimetry exam*.mp. | 0 | Advanced |
| 187 | campimetr*.mp. | 17 | Advanced |
| 188 | perimetr*.mp. | 1147 | Advanced |
| 189 | tangent screen exam*.mp. | 1 | Advanced |
| 190 | auditory perception.mp. | 1064 | Advanced |
| 191 | auditory processing.mp. | 299 | Advanced |
| 192 | (auditory adj2 localization*).mp. | 14 | Advanced |
| 193 | (sound adj2 localization*).mp. | 141 | Advanced |
| 194 | spatial hearing.mp. | 31 | Advanced |
| 195 | or/1-194 | 59125 | Advanced |
| 196 | Virtual Reality/ | 1162 | Advanced |
| 197 | (virtual adj3 realit*).mp. | 7374 | Advanced |
| 198 | (augmented adj3 realit*).mp. | 624 | Advanced |
| 199 | VRET.mp. | 108 | Advanced |
| 200 | iVR.mp. | 739 | Advanced |
| 201 | iVRs.mp. | 207 | Advanced |
| 202 | iVE.mp. | 165 | Advanced |
| 203 | iVEs.mp. | 32 | Advanced |
| 204 | (immers* adj2 environment*).mp. | 171 | Advanced |
| 205 | samsung gear.mp. | 17 | Advanced |
| 206 | oculus rift.mp. | 66 | Advanced |
| 207 | HCD vive.mp. | 0 | Advanced |
| 208 | hololens.mp. | 65 | Advanced |
| 209 | gear VR.mp. | 12 | Advanced |
| 210 | head-mount*.mp. | 581 | Advanced |
| 211 | (head adj3 gear*).mp. | 20 | Advanced |
| 212 | HMD.mp. | 241 | Advanced |
| 213 | Gen2-VR.mp. | 0 | Advanced |
| 214 | (google adj2 daydream).mp. | 2 | Advanced |
| 215 | (google adj2 cardboard).mp. | 13 | Advanced |
| 216 | PlayStation.mp. | 39 | Advanced |
| 217 | (ReTrak adj2 Utopia).mp. | 0 | Advanced |
| 218 | NOON VR.mp. | 1 | Advanced |
| 219 | Freefly VR.mp. | 0 | Advanced |
| 220 | Homido.mp. | 0 | Advanced |
| 221 | iWear.mp. | 0 | Advanced |
| 222 | (mixed adj2 realit*).mp. | 162 | Advanced |
| 223 | (simulat* adj2 environment*).mp. | 486 | Advanced |
| 224 | (virtual adj2 environment*).mp. | 1064 | Advanced |
| 225 | Video Games/ | 1179 | Advanced |
| 226 | (computer adj2 game*).mp. | 722 | Advanced |
| 227 | (video adj2 game*).mp. | 2626 | Advanced |
| 228 | gaming.mp. | 1128 | Advanced |
| 229 | user-computer interface/ | 1523 | Advanced |
| 230 | (user adj2 interface*).mp. | 1886 | Advanced |
| 231 | (virtual adj2 interface*).mp. | 24 | Advanced |
| 232 | Oculus Quest.mp. | 57 | Advanced |
| 233 | Meta Quest.mp. | 16 | Advanced |
| 234 | Metaverse.mp. | 29 | Advanced |
| 235 | (simulate* adj2 realit*).mp. | 17 | Advanced |
| 236 | (tethered adj2 headset*).mp. | 0 | Advanced |
| 237 | Serious game.mp. | 340 | Advanced |
| 238 | or/196-237 | 14372 | Advanced |
| 239 | 195 and 238 | 657 | Advanced |
| 240 | remove duplicates from 239 | 645 | Advanced |
| 241 | ("202308$" or "202309$" or "202310$" or "202311$" or "202312$" or "2024$").up. | 513499 | Advanced |
| 242 | 240 and 241 | 188 | Advanced |

EBM Reviews - Cochrane Database of Systematic Reviews <2005 to August 14, 2024>

| **#** | **Searches** | **Results** | **Type** |
| --- | --- | --- | --- |
| 1 | (eye adj2 disease*).ti,ab. | 16 | Advanced |
| 2 | (eye adj2 disorder*).ti,ab. | 3 | Advanced |
| 3 | Albinism.ti,ab. | 0 | Advanced |
| 4 | (Chediak adj2 higashi syndrome).ti,ab. | 0 | Advanced |
| 5 | (Congenital adj2 amauros?s).ti,ab. | 0 | Advanced |
| 6 | dysgenesis neuroepithelialis retinae.ti,ab. | 0 | Advanced |
| 7 | hereditary epithelial dysplasia of retina.ti,ab. | 0 | Advanced |
| 8 | hereditary retinal aplasia.ti,ab. | 0 | Advanced |
| 9 | heredoretinopathia congenitalis.ti,ab. | 0 | Advanced |
| 10 | leber abiotroph*.ti,ab. | 0 | Advanced |
| 11 | (leber* adj2 amauros?s).ti,ab. | 0 | Advanced |
| 12 | leber congenital tapetoretinal degeneration.ti,ab. | 0 | Advanced |
| 13 | hereditary optic atroph*.ti,ab. | 0 | Advanced |
| 14 | (optic adj2 hypoplasia*).ti,ab. | 0 | Advanced |
| 15 | (retinal adj2 degeneration*).ti,ab. | 1 | Advanced |
| 16 | (retinal adj2 dysplasia*).ti,ab. | 0 | Advanced |
| 17 | pigmentary retinopathy*.ti,ab. | 0 | Advanced |
| 18 | retinitis pigmentosa.ti,ab. | 1 | Advanced |
| 19 | tapetoretinal degeneration*.ti,ab. | 0 | Advanced |
| 20 | fundus flavimaculatus.ti,ab. | 0 | Advanced |
| 21 | macular dystrophy with flecks.ti,ab. | 0 | Advanced |
| 22 | stargardt disease.ti,ab. | 0 | Advanced |
| 23 | cod md syndrome*.ti,ab. | 0 | Advanced |
| 24 | cerebromuscular dystrophy.ti,ab. | 0 | Advanced |
| 25 | cerebroocular dysplasia muscular dystrophy syndrome.ti,ab. | 0 | Advanced |
| 26 | chemke syndrome.ti,ab. | 0 | Advanced |
| 27 | congenital muscular dystrophy dystroglycanopathy.ti,ab. | 0 | Advanced |
| 28 | walker-warburg syndrome*.ti,ab. | 0 | Advanced |
| 29 | fukuyama cmd.ti,ab. | 0 | Advanced |
| 30 | (fukuyama adj3 muscular dystrophy).ti,ab. | 0 | Advanced |
| 31 | fukuyama syndrome.ti,ab. | 0 | Advanced |
| 32 | hard syndrome*.ti,ab. | 0 | Advanced |
| 33 | lgmd2k.ti,ab. | 0 | Advanced |
| 34 | mddga1.ti,ab. | 0 | Advanced |
| 35 | meb syndrome.ti,ab. | 0 | Advanced |
| 36 | muscle-eye-brain syndrome.ti,ab. | 0 | Advanced |
| 37 | muscle eye brain disease*.ti,ab. | 0 | Advanced |
| 38 | muscular dystrophy due to defective glycosylation of dystroglycan 4a.ti,ab. | 0 | Advanced |
| 39 | muscular dystrophy-dystroglycanopathy.ti,ab. | 0 | Advanced |
| 40 | pagon syndrome*.ti,ab. | 0 | Advanced |
| 41 | warburg syndrome.ti,ab. | 0 | Advanced |
| 42 | alpha dystroglycanopathies.ti,ab. | 0 | Advanced |
| 43 | congenital mesodermal dysmorphodystroph*.ti,ab. | 0 | Advanced |
| 44 | gems.ti,ab. | 1 | Advanced |
| 45 | glaucoma-lens ectopia-microspherophakia-stiffness-shortness syndrome.ti,ab. | 0 | Advanced |
| 46 | (marchesani adj2 syndrome*).ti,ab. | 0 | Advanced |
| 47 | spherophakia brachymorphia syndrome*.ti,ab. | 0 | Advanced |
| 48 | cataract*.ti,ab. | 78 | Advanced |
| 49 | lens opacity*.ti,ab. | 2 | Advanced |
| 50 | pseudoaphakia*.ti,ab. | 0 | Advanced |
| 51 | (lens adj2 cloud*).ti,ab. | 0 | Advanced |
| 52 | glaucoma*.ti,ab. | 62 | Advanced |
| 53 | increased intraocular pressure.ti,ab. | 4 | Advanced |
| 54 | ocular hypertension.ti,ab. | 12 | Advanced |
| 55 | optic neuropathy.ti,ab. | 15 | Advanced |
| 56 | brown* tendon sheath syndrome*.ti,ab. | 0 | Advanced |
| 57 | conjugate gaze spasm*.ti,ab. | 0 | Advanced |
| 58 | convergence excess*.ti,ab. | 0 | Advanced |
| 59 | convergence insufficienc*.ti,ab. | 1 | Advanced |
| 60 | cyclophoria*.ti,ab. | 0 | Advanced |
| 61 | eye motility disorder*.ti,ab. | 0 | Advanced |
| 62 | eye movement disorder*.ti,ab. | 3 | Advanced |
| 63 | internuclear ophthalmoplegia*.ti,ab. | 0 | Advanced |
| 64 | ocular motility disorder*.ti,ab. | 0 | Advanced |
| 65 | ocular torticollis.ti,ab. | 1 | Advanced |
| 66 | opsoclonus.ti,ab. | 0 | Advanced |
| 67 | parinaud* syndrome*.ti,ab. | 0 | Advanced |
| 68 | paroxysmal ocular dyskinesia*.ti,ab. | 0 | Advanced |
| 69 | pseudoophthalmoplegia*.ti,ab. | 0 | Advanced |
| 70 | skew deviation*.ti,ab. | 0 | Advanced |
| 71 | smooth pursuit deficienc*.ti,ab. | 0 | Advanced |
| 72 | spasm of conjugate gaze.ti,ab. | 0 | Advanced |
| 73 | tendon sheath syndrome of brown.ti,ab. | 0 | Advanced |
| 74 | fisher syndrome.ti,ab. | 1 | Advanced |
| 75 | miller fisher.ti,ab. | 0 | Advanced |
| 76 | (ophthalmoplegia, ataxia and areflexia syndrome).ti,ab. | 0 | Advanced |
| 77 | nystagmus.ti,ab. | 8 | Advanced |
| 78 | involuntary eye movement*.ti,ab. | 1 | Advanced |
| 79 | Jerky eye movement*.ti,ab. | 0 | Advanced |
| 80 | cranial nerve iii disease*.ti,ab. | 0 | Advanced |
| 81 | oculomotor nerve disease*.ti,ab. | 0 | Advanced |
| 82 | oculomotor nerve disorder*.ti,ab. | 0 | Advanced |
| 83 | oculomotor nerve pals*.ti,ab. | 0 | Advanced |
| 84 | oculomotor nerve paralys?s.ti,ab. | 0 | Advanced |
| 85 | oculomotor neuropath*.ti,ab. | 0 | Advanced |
| 86 | third cranial nerve disease*.ti,ab. | 0 | Advanced |
| 87 | third nerve pals*.ti,ab. | 0 | Advanced |
| 88 | third nerve paralysis.ti,ab. | 1 | Advanced |
| 89 | ophthalmoplegia*.ti,ab. | 1 | Advanced |
| 90 | oculomotor paralysis.ti,ab. | 0 | Advanced |
| 91 | ophthalmopares?s.ti,ab. | 0 | Advanced |
| 92 | dancing eyes.ti,ab. | 0 | Advanced |
| 93 | myoclonic encephalopathy*.ti,ab. | 0 | Advanced |
| 94 | kinsbourne syndrome.ti,ab. | 0 | Advanced |
| 95 | opsoclonus myoclonus.ti,ab. | 0 | Advanced |
| 96 | strabismus.ti,ab. | 14 | Advanced |
| 97 | dissociated horizontal deviation*.ti,ab. | 0 | Advanced |
| 98 | dissociated vertical deviation*.ti,ab. | 2 | Advanced |
| 99 | heterophoria*.ti,ab. | 0 | Advanced |
| 100 | heterotropia*.ti,ab. | 0 | Advanced |
| 101 | hypertropia*.ti,ab. | 1 | Advanced |
| 102 | phoria*.ti,ab. | 0 | Advanced |
| 103 | squint*.ti,ab. | 3 | Advanced |
| 104 | crossed eye*.ti,ab. | 0 | Advanced |
| 105 | tolosa hunt syndrome*.ti,ab. | 0 | Advanced |
| 106 | Ametropia*.ti,ab. | 0 | Advanced |
| 107 | refractive disorder*.ti,ab. | 0 | Advanced |
| 108 | refractive error*.ti,ab. | 20 | Advanced |
| 109 | aniseikonia.ti,ab. | 0 | Advanced |
| 110 | anisometropia.ti,ab. | 1 | Advanced |
| 111 | astigmatism*.ti,ab. | 13 | Advanced |
| 112 | corneal wavefront aberration*.ti,ab. | 0 | Advanced |
| 113 | farsighted*.ti,ab. | 0 | Advanced |
| 114 | longsighted*.ti,ab. | 0 | Advanced |
| 115 | hypermetropia*.ti,ab. | 1 | Advanced |
| 116 | hyperopia*.ti,ab. | 3 | Advanced |
| 117 | myopia*.ti,ab. | 14 | Advanced |
| 118 | nearsighted*.ti,ab. | 2 | Advanced |
| 119 | shortsighted*.ti,ab. | 0 | Advanced |
| 120 | presbyopia*.ti,ab. | 3 | Advanced |
| 121 | (retinal adj2 disease*).ti,ab. | 1 | Advanced |
| 122 | (cone adj2 dystroph*).ti,ab. | 0 | Advanced |
| 123 | diabetic retinopath*.ti,ab. | 31 | Advanced |
| 124 | hypertensive retinopath*.ti,ab. | 0 | Advanced |
| 125 | diabetic eye disease*.ti,ab. | 0 | Advanced |
| 126 | diabetic macular edema.ti,ab. | 2 | Advanced |
| 127 | blindness.ti,ab. | 72 | Advanced |
| 128 | hemeralopia*.ti,ab. | 0 | Advanced |
| 129 | macropsia*.ti,ab. | 0 | Advanced |
| 130 | metamorphopsia*.ti,ab. | 2 | Advanced |
| 131 | micropsia*.ti,ab. | 0 | Advanced |
| 132 | vision disabilit*.ti,ab. | 0 | Advanced |
| 133 | vision disorder*.ti,ab. | 1 | Advanced |
| 134 | vision impairment*.ti,ab. | 3 | Advanced |
| 135 | visual disorder*.ti,ab. | 0 | Advanced |
| 136 | visual impairment*.ti,ab. | 28 | Advanced |
| 137 | visual disabilit*.ti,ab. | 3 | Advanced |
| 138 | amblyopia*.ti,ab. | 14 | Advanced |
| 139 | lazy eye*.ti,ab. | 0 | Advanced |
| 140 | amauros?s.ti,ab. | 1 | Advanced |
| 141 | sudden visual loss*.ti,ab. | 0 | Advanced |
| 142 | achromatopsia*.ti,ab. | 0 | Advanced |
| 143 | colo?r vision defect*.ti,ab. | 0 | Advanced |
| 144 | colo?r vision deficienc*.ti,ab. | 0 | Advanced |
| 145 | colo?r vision impairment*.ti,ab. | 0 | Advanced |
| 146 | colo?r perception deficienc*.ti,ab. | 0 | Advanced |
| 147 | deutan defect.ti,ab. | 0 | Advanced |
| 148 | monochromatopsia.ti,ab. | 0 | Advanced |
| 149 | protan defect.ti,ab. | 0 | Advanced |
| 150 | tritan defect.ti,ab. | 0 | Advanced |
| 151 | diplopia*.ti,ab. | 14 | Advanced |
| 152 | double vision.ti,ab. | 4 | Advanced |
| 153 | polyopsia*.ti,ab. | 0 | Advanced |
| 154 | nyctalopia.ti,ab. | 1 | Advanced |
| 155 | light sensitivit*.ti,ab. | 1 | Advanced |
| 156 | photophobia*.ti,ab. | 10 | Advanced |
| 157 | scotoma*.ti,ab. | 1 | Advanced |
| 158 | retinocochleocerebral vasculopath*.ti,ab. | 0 | Advanced |
| 159 | susac* syndrome.ti,ab. | 0 | Advanced |
| 160 | diminished vision.ti,ab. | 0 | Advanced |
| 161 | reduced vision.ti,ab. | 0 | Advanced |
| 162 | subnormal vision.ti,ab. | 0 | Advanced |
| 163 | sub-normal vision.ti,ab. | 0 | Advanced |
| 164 | low vision.ti,ab. | 7 | Advanced |
| 165 | macular degeneration*.ti,ab. | 32 | Advanced |
| 166 | maculopath*.ti,ab. | 0 | Advanced |
| 167 | macular dystroph*.ti,ab. | 0 | Advanced |
| 168 | macular disorder*.ti,ab. | 0 | Advanced |
| 169 | retinal degeneration*.ti,ab. | 0 | Advanced |
| 170 | retinal degenerative disease*.ti,ab. | 0 | Advanced |
| 171 | visual field*.ti,ab. | 36 | Advanced |
| 172 | vision test*.ti,ab. | 2 | Advanced |
| 173 | colo?r perception test*.ti,ab. | 0 | Advanced |
| 174 | ocular refraction*.ti,ab. | 0 | Advanced |
| 175 | vision screening.ti,ab. | 5 | Advanced |
| 176 | visual acuit*.ti,ab. | 146 | Advanced |
| 177 | visual contrast sensitivity*.ti,ab. | 0 | Advanced |
| 178 | emmetropia*.ti,ab. | 0 | Advanced |
| 179 | automated perimetry exam*.ti,ab. | 0 | Advanced |
| 180 | campimetr*.ti,ab. | 0 | Advanced |
| 181 | perimetr*.ti,ab. | 1 | Advanced |
| 182 | tangent screen exam*.ti,ab. | 0 | Advanced |
| 183 | auditory perception.ti,ab. | 0 | Advanced |
| 184 | auditory processing.ti,ab. | 1 | Advanced |
| 185 | (auditory adj2 localization*).ti,ab. | 0 | Advanced |
| 186 | (sound adj2 localization*).ti,ab. | 0 | Advanced |
| 187 | spatial hearing.ti,ab. | 0 | Advanced |
| 188 | or/1-187 | 333 | Advanced |
| 189 | (virtual adj3 realit*).ti,ab. | 18 | Advanced |
| 190 | (augmented adj3 realit*).ti,ab. | 0 | Advanced |
| 191 | VRET.ti,ab. | 0 | Advanced |
| 192 | iVR.ti,ab. | 1 | Advanced |
| 193 | iVRs.ti,ab. | 0 | Advanced |
| 194 | iVE.ti,ab. | 0 | Advanced |
| 195 | iVEs.ti,ab. | 0 | Advanced |
| 196 | (immers* adj2 environment*).ti,ab. | 0 | Advanced |
| 197 | samsung gear.ti,ab. | 0 | Advanced |
| 198 | oculus rift.ti,ab. | 0 | Advanced |
| 199 | HCD vive.ti,ab. | 0 | Advanced |
| 200 | hololens.ti,ab. | 0 | Advanced |
| 201 | gear VR.ti,ab. | 0 | Advanced |
| 202 | head-mount*.ti,ab. | 1 | Advanced |
| 203 | (head adj3 gear*).ti,ab. | 0 | Advanced |
| 204 | HMD.ti,ab. | 1 | Advanced |
| 205 | Gen2-VR.ti,ab. | 0 | Advanced |
| 206 | (google adj2 daydream).ti,ab. | 0 | Advanced |
| 207 | (google adj2 cardboard).ti,ab. | 0 | Advanced |
| 208 | PlayStation.ti,ab. | 0 | Advanced |
| 209 | (ReTrak adj2 Utopia).ti,ab. | 0 | Advanced |
| 210 | NOON VR.ti,ab. | 0 | Advanced |
| 211 | Freefly VR.ti,ab. | 0 | Advanced |
| 212 | Homido.ti,ab. | 0 | Advanced |
| 213 | iWear.ti,ab. | 0 | Advanced |
| 214 | (mixed adj2 realit*).ti,ab. | 0 | Advanced |
| 215 | (simulat* adj2 environment*).ti,ab. | 2 | Advanced |
| 216 | (virtual adj2 environment*).ti,ab. | 3 | Advanced |
| 217 | (computer adj2 game*).ti,ab. | 6 | Advanced |
| 218 | (video adj2 game*).ti,ab. | 2 | Advanced |
| 219 | gaming.ti,ab. | 7 | Advanced |
| 220 | (user adj2 interface*).ti,ab. | 1 | Advanced |
| 221 | (virtual adj2 interface*).ti,ab. | 0 | Advanced |
| 222 | Oculus Quest.ti,ab. | 0 | Advanced |
| 223 | Meta Quest.ti,ab. | 0 | Advanced |
| 224 | Metaverse.ti,ab. | 0 | Advanced |
| 225 | (simulate* adj2 realit*).ti,ab. | 0 | Advanced |
| 226 | (tethered adj2 headset*).ti,ab. | 0 | Advanced |
| 227 | Serious game.ti,ab. | 0 | Advanced |
| 228 | or/189-227 | 30 | Advanced |
| 229 | 188 and 228 | 3 | Advanced |
| 230 | ("202308$" or "202309$" or "202310$" or "202311$" or "202312$" or "2024$").up. | 3172 | Advanced |
| 231 | 229 and 230 | 1 | Advanced |

*Consider, if feasible to do so, reporting the number of records identified from each database or register searched (rather than the total number across all databases/registers).

**If automation tools were used, indicate how many records were excluded by a human and how many were excluded by automation tools.

*From:*  Page MJ, McKenzie JE, Bossuyt PM, Boutron I, Hoffmann TC, Mulrow CD, et al. The PRISMA 2020 statement: an updated guideline for reporting systematic reviews. BMJ 2021;372:n71. doi: 10.1136/bmj.n71

For more information, visit: <http://www.prisma-statement.org/>
